# The long-term epidemiological impacts of doxycycline post-exposure prophylaxis and vaccination against multidrug-resistant *Neisseria gonorrhoeae*: a mathematical modelling study

**DOI:** 10.64898/2026.07.28.26359123

**Authors:** Zihao Wang, Dariya Nikitin, George Young, Matan Yechezkel, Liang En Wee, Martin T.W. Chio, Lin Geng, Rayner Kay Jin Tan, Yi Wang, David N. Fisman, Joseph A. Lewnard, Lilith K. Whittles, Jue Tao Lim

## Abstract

Doxycycline post-exposure prophylaxis (doxy-PEP) represents major advancements in sexually transmitted infection prevention for men who have sex with men (MSM), yet the joint long-term population-level impact on *Neisseria gonorrhoeae* transmission dynamics and antimicrobial resistance (AMR) together with a moderately effective vaccine remains uncertain. Here, we developed a deterministic transmission model calibrated to empirical surveillance data from England to evaluate the 15-year epidemiological interactions of implementing doxy-PEP and vaccination at the population level. The model incorporates the tetracycline- and ceftriaxone-susceptible, tetracycline-resistant (Tet-R), ceftriaxone-resistant (Cef-R), and dual-resistant (Dual-R) strains. Our simulations suggest that the unmitigated deployment of doxy-PEP provides only modest reductions in overall gonorrhoea burden, yielding a net programmatic efficiency of 0.072 (95% CrI: 0.018 – 0.23) averted infections per enrolment over the 15-year horizon. Although doxy-PEP reduces tetracycline- and ceftriaxone-susceptible infections, it consistently selects for Tet-R lineages. Conversely, standalone vaccination yields substantially greater epidemiological benefit, averting 0.73 (95% CrI: 0.17 – 1.63) overall infections per enrolment. When deployed in tandem, the dual intervention strategy achieves the greatest overall effectiveness to 0.83 (95% CrI: 0.22 – 1.71) averted infections per enrolment while mitigating the Tet-R selection pressure observed under standalone doxy-PEP. Long-term strain dynamics were found to be governed by background frontline ceftriaxone treatment failure rates rather than intervention uptake. Under a scenario of compromised ceftriaxone efficacy (20% treatment failure), doxy-PEP is projected to favour expansion of Dual-R strain, yielding 510 (95% CrI: 38 –258600) excess Dual-R infections across the 15-year horizon (146.18% [95% CrI: 31.30 – 920.86%] cumulative increase) and triggering sustained transmission exceeding the no-intervention baseline. Conversely, vaccination standalone or combined strategies consistently suppress overall transmission. By year 15, annual Cef-R and Dual-R infections are restricted to ≤ 19 infections under both vaccine-inclusive approaches. These findings suggest that the public health utility of antibiotic prophylaxis is heavily contingent upon the preserved efficacy of frontline therapeutics. Our work demonstrates that single-agent prophylaxis risks localized containment of some pathogens at the cost of driving multidrug-resistant selection in others and underscores the necessity of integrating non-selective tools like vaccines to manage *N. gonorrhoeae*.

**One Sentence Summary:** Combining vaccination with doxycycline prophylaxis improves gonorrhoea control while limiting antimicrobial resistance.

## Introduction

Doxycycline, a tetracycline-class broad-spectrum antibiotic, can be used as post-exposure prophylaxis (doxy- PEP) to prevent syphilis and other bacterial sexually transmitted infections among men who have sex with men (MSM) [1], [2]. However, its large-scale deployment has intensified concerns regarding collateral selection pressures on antimicrobial resistance (AMR) in *Neisseria gonorrhoeae* [3], [4], [5], [6], [7], [8]. Although resistance determinants for doxycycline [9], [10] and extended-spectrum cephalosporins (such as ceftriaxone, the global standard-of-care monotherapy) [11] arise through independent genetic pathways, they can become epidemiologically linked via co-selection [3], [5], [12]. Notably, ceftriaxone-resistant, multidrug-resistant *N. gonorrhoeae* lineages have already emerged globally, with documented transmission in jurisdictions including Singapore [13] and England [14]. While novel antibiotics are in development (e.g., oral gepotidacin [15] and zoliflodacin [16]), their clinical utility remains constrained. For instance, though the novel type II topoisomerase inhibitor zoliflodacin demonstrates robust *in vitro* activity against multidrug-resistant *N. gonorrhoeae* [16], [17], recent clinical trial data indicate notable therapeutic vulnerabilities. Oral zoliflodacin effectively clears uncomplicated urogenital and rectal gonococcal infections, yet exhibits significantly lower efficacy in pharyngeal compartments, ultimately remaining clinically inferior to ceftriaxone [18]. Consequently, given the limitations of the therapeutics pipeline, evaluating how public health interventions can preserve existing ceftriaxone efficacy is crucial.

Concurrently, studies on the four-component meningococcal serogroup B (4CMenB) vaccine has suggested partial cross-protection against *N. gonorrhoeae* mediated by highly conserved shared surface antigens [19], [20]. Furthermore, while observational epidemiological data suggest that 4CMenB deployment reduces gonorrhoea incidence by 30% to 40% [21], recent randomized controlled trials have reported contrasting outcomes, finding no significant efficacy against gonococcal infection [22], [23], [24]. These discordant findings highlight the need for cautious implementation and further data from ongoing trials evaluating meningococcal vaccines across diverse populations, while underscoring the urgent necessity for a gonorrhoea-specific vaccine. Nonetheless, incorporating a vaccine with even moderate protective efficacy into public health frameworks may be a crucial strategy [19]. By potentially suppressing overall transmission dynamics and decreasing baseline incidence, vaccination can reduce the net volume of clinical cases. This down-regulation subsequently diminishes population-level antibiotic consumption, weakening the selective pressure driving AMR and attenuating expansion of resistant lineages.

England represents a particularly important setting in which to evaluate these competing evolutionary and public health pressures. The UK Health Security Agency (UKHSA) has recently recommended targeted deployment of both doxy-PEP and the 4CMenB vaccine for eligible MSM, making England one of the first settings to consider simultaneous implementation of antibiotic prophylaxis and vaccination within a national sexual health programme [21], [25]. At the same time, gonococcal tetracycline resistance is already common among MSM in England, with 68.5% of isolates demonstrating tetracycline resistance (MIC^1^ > 1mg/L) in 2022 and 30.1% exhibiting high-level resistance (MIC ≥ 8mg/L) in 2024 [26]. This relatively high, but not yet saturated, prevalence creates a unique epidemiological environment in which doxy-PEP may provide short-term protection while simultaneously exerting substantial selective pressure for further resistance. Consequently, England provides an ideal natural policy setting for evaluating whether vaccination can offset these evolutionary consequences.

Although previous mathematical modelling has established that antibiotic exposure accelerates the selection of AMR [27], [28], [29], whereas vaccination can suppress resistant *N. gonorrhoeae* lineages [30], [31], no study has evaluated the long-term interaction between these two interventions within the policy context currently being implemented in England. In particular, it remains unknown whether introducing doxy-PEP into a population where tetracycline resistance is already common will continue to provide meaningful population-level benefits or instead accelerate competitive strain replacement towards resistant lineages. Likewise, it remains unclear whether concurrent vaccination can sufficiently reduce transmission to offset these evolutionary pressures.

1 Minimum Inhibitory Concentration

To address these policy-relevant uncertainties, we developed a deterministic, strain-structured transmission model to project the long-term population-level AMR dynamics of *N. gonorrhoeae* over a 15-year horizon within the epidemiological context of England and thus to quantify both the long-term programmatic efficiency and the evolutionary consequences of doxy-PEP and vaccination by jointly evaluating infections averted per enrolment and strain-specific resistance dynamics. The model characterises the transmission of four *N. gonorrhoeae* strains, namely, tetracycline- and ceftriaxone-susceptible (hereafter referred to as the baseline susceptible), tetracycline-resistant (Tet-R), ceftriaxone-resistant (Cef-R), and dual-resistant (Dual-R) strains. Moreover, the model structure accounts for behavioural risk heterogeneity and differential intervention uptake across four distinct strata: individuals receiving neither intervention, doxy-PEP standalone intervention, vaccination standalone intervention, or combined interventions. We calibrated the model within a Bayesian framework using longitudinal surveillance data from England. Through counterfactual simulations compared against a no-intervention baseline, we quantified the intersection of dual selective pressures - doxycycline-induced selection and clinical treatment failure - under varying coverage thresholds of doxy-PEP for syphilis and vaccination for gonorrhoea. Crucially, this framework evaluates the extent to which population-level doxy-PEP deployment among MSM might inadvertently amplify the transmission of multidrug-resistant *N. gonorrhoeae* against a backdrop of relatively high but not yet saturated baseline tetracycline resistance [32]. Moreover, we characterize the capacity of a moderate-efficacy vaccine to attenuate the emergence of dual resistance and mitigate the evolutionary selective pressures introduced by expanded doxy-PEP for syphilis utility.

## Methods

A comprehensive description of the mathematical modelling workflow, comprising the model specification, parameterization, calibration, and intervention simulations, is detailed in the Supplementary Information Section 2 and Section 3. To ensure methodological transparency and reproducibility, all structural parameters, state variables, and initial conditions are explicitly codified alongside their respective point estimates, probability distributions, and primary empirical data sources. Source code to reproduce all our experiments, figures, and analysis is publicly available at the GitHub repository: https://github.com/killingbear999/amr_gonorrhoea.

### 1. Data

England serves as an ideal epidemiological setting for evaluating the evolutionary dynamics of *N. gonorrhoeae* AMR under dual intervention pressures. The jurisdiction maintains world-class, comprehensive national surveillance registries, specifically Genitourinary Medicine Clinic Activity Dataset (GUMCAD) [33] and Gonococcal Resistance to Antimicrobials Surveillance Programme (GRASP) [34], which provide high-resolution, longitudinal data on sexually transmitted infection (STI) testing volumes, clinical presentations, and strain-specific resistance profiles. Furthermore, transmission in the English MSM population is characterized by a relatively high but not yet saturated baseline prevalence of tetracycline resistance [32] and a complete absence of sustained and domestically acquired ceftriaxone-resistant *N. gonorrhoeae* clusters to date [14], but there are rising pressures on frontline ceftriaxone susceptibility reported in England since 2021 [26]. This context has the epidemiological data required for model calibration and evaluating targeted public health interventions. See Supplementary Information Section 2 for full details.

### 2. Interventions

To evaluate the long-term epidemiological impacts of widespread doxy-PEP and vaccination rollout against *N. gonorrhoeae*, we simulated two distinct interventions across a 15-year horizon (2027-2041), implemented both as standalone and combined strategies against a baseline counterfactual of standard care. The first intervention, doxy-PEP (indicated primarily for syphilis prevention but providing secondary, partial protection against *N. gonorrhoeae*), was characterized as an antibiotic-based, systemic chemoprophylactic strategy targeting MSM following condomless sexual exposure. Mechanistically, it was modelled as an agent-selective filter that exerts bactericidal efficacy against baseline susceptible strains but exhibits a complete ineffectiveness as prophylaxis against pre-existing Tet-R or Dual-R determinants. The second intervention, a moderately effective vaccine, was evaluated as an evolutionary-neutral, transmission-blocking barrier that reduces host susceptibility uniformly across all bacterial strains regardless of their antimicrobial resistance profiles. For the combined intervention, individuals receiving both doxy-PEP and vaccination simultaneously experienced the complementary effects of both interventions, whereby vaccination reduced susceptibility to infection across all strains, while doxy-PEP provided additional post-exposure protection against doxycycline-susceptible strains only. Given that doxy-PEP and vaccination have only recently been introduced in England, empirical data on population-level uptake remain limited. Thus, the baseline uptake rate for both interventions is assumed to be 66% in the main analysis, reflecting high empirical willingness to adopt prophylaxis (> 60%) documented in recent behavioural surveys from Ireland [35] and Australia [36], [37]. Simulations were evaluated over a 15-year horizon to allow sufficient time for population-level strain dynamics and selective pressures to reach steady-state conditions. This long-term analysis explicitly incorporates key parameters governing the intervention, including vaccine immunity waning and doxy-PEP discontinuation. Simulations of the interventions were thereafter subject to sensitivity analyses to evaluate the effect of varying background ceftriaxone treatment failure rates and intervention uptake rates on intervention effectiveness.

### 3. Model structure

To quantify the population-level impact of doxy-PEP prevention on the transmission dynamics of resistant *N. gonorrhoeae* among MSM, we constructed a deterministic, strain-specific compartmental model (see Fig. 1, detailed in Supplementary Information Section 3). The framework also evaluates the capacity of a moderate-efficacy vaccine to mitigate AMR emergence and countervail the selective pressures introduced by expanded prophylactic antibiotic use. To maintain analytical tractability and isolate the precise evolutionary vectors of interest, our modelling framework focused on the four strains directly implicated by frontline therapy and doxy- PEP deployment: baseline susceptible, Tet-R, Cef-R, and Dual-R [29]. Consequently, all other historically resistant lineages, including those demonstrating resistance to azithromycin, cefixime, ciprofloxacin, or penicillin, were aggregated into a single reference “baseline susceptible” compartment. This consolidation helps focus on the specific selective pressures exerted by the simulated interventions and is justified because these non-target resistance profiles do not share direct mechanistic cross-resistance with ceftriaxone or doxycycline [38]. Moreover, to reproduce the low endemic prevalence of Cef-R and Dual-R strains observed in England surveillance data, each resistant lineage is assigned a strain-specific relative fitness parameter ( *f* ≤ 1 ), representing the reduction in transmissibility associated with antimicrobial resistance in the absence of selective antibiotic pressure. These fitness penalties counterbalance the continual generation of resistant strains through treatment and prophylactic selection, allowing resistant lineages to persist at low endemic frequencies while remaining capable of expansion when selective pressures increase.

**Fig. 1:**
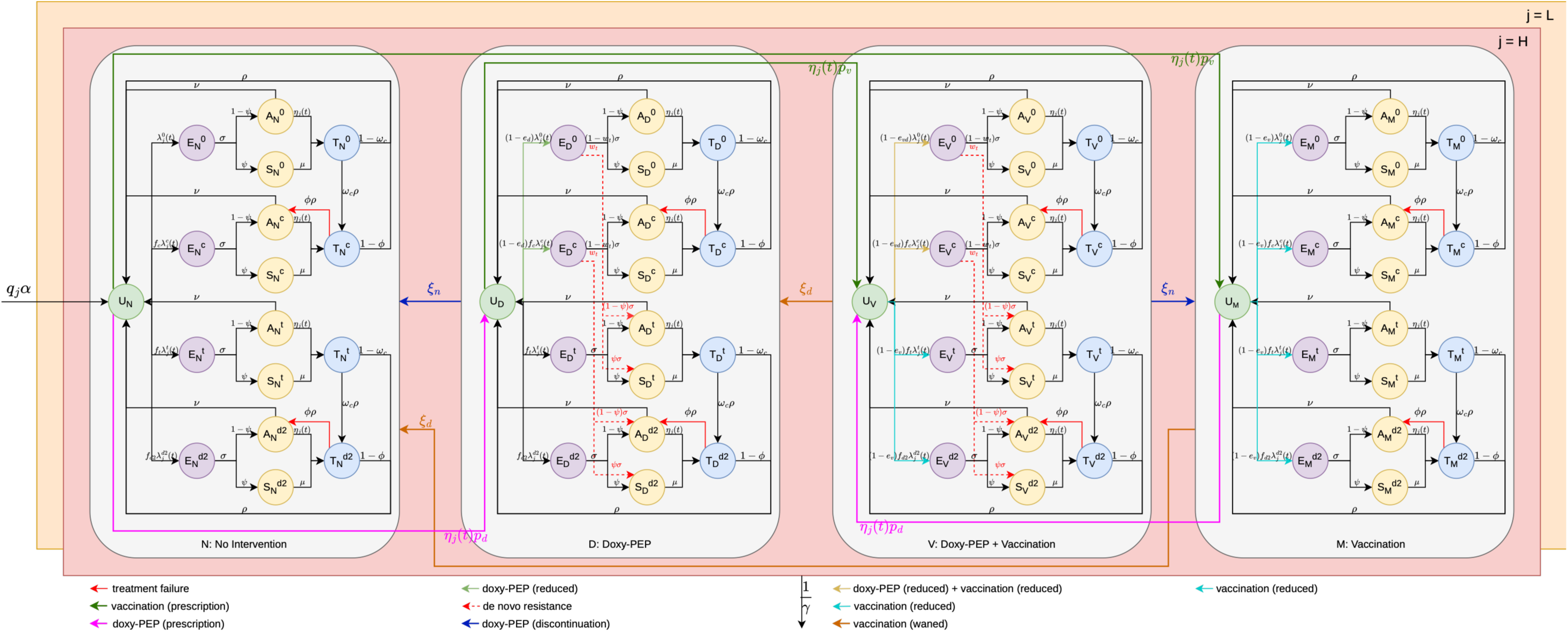
Compartmental structure and transmission architecture of the strain-stratified *N. gonorrhoeae* model under dual intervention pressures. The schematic illustrates the transmission dynamics and evolutionary pathways of *N. gonorrhoeae* under targeted doxy-PEP and vaccination. The target MSM population is stratified into low-activity (orange) and high-activity (red) sexual behaviour classes. Both classes share identical internal compartmental structures but possess distinct partner-change and transmission transition rates; for visual clarity, only inter-compartmental flows for the high-activity cohort (upper layer) are explicitly plotted. Demographic renewal is maintained by the continuous entry of individuals into the sexually active population as uninfected, fully susceptible dynamics (*U*) within the non-intervention stratum (*N*) at a rate *α*, balanced by a constant background attrition rate across all states due to aging or exit from the active network at a rate 1/*γ*. The population is split into four distinct sub-populations based on coverage and uptake status: non-intervention (*N*), doxy-PEP standalone intervention (*D*), vaccination standalone intervention (*V*), and combined dual-interventions (*M*). Within each stratum (*i* ∈ {*N*, *D*, *V*, *M*}), clinical progression follows an extended Susceptible-Exposed-Infectious-Treated framework: uninfected (*U*) individuals transition to exposed (*E*) driven by the time-varying strain-specific force of infection *λ* and a corresponding relative fitness *f*, progressing subsequently at a rate *σ* to either asymptomatic (*A*) or symptomatic (*S*) acute infectious states with a probability 1 − *ψ* and *ψ*, respectively. Screened individuals including both symptomatic and asymptomatic hosts enter the treated compartment (*T*) prior to recovery at rates *μ* and *η* respectively, while untreated asymptomatic infections can undergo natural clearance (*A* → *U*) at a rate *ν*. Following successful therapeutic intervention, treated individuals return to the susceptible state (*T* → *U*) at a rate *ρ* while individuals with Cef-R and Dual-R infections experiencing frontline therapeutic treatment failure with a probability of *φ* revert to the actively infectious asymptomatic compartment (*T* → *A*; red arrows) at a rate *ρ*, establishing a persistent reservoir of transmissible infection. For analytical simplicity, intervention initiation is restricted to the uninfected (*U*) state. Infections within each sub-population are disaggregated across four co-circulating gonococcal strains characterized by distinct resistance profiles (superscript *k*): baseline susceptible (0), ceftriaxone-resistant (*c*), tetracycline-resistant (*t*), and dual-resistant (*d*2). Ceftriaxone is modelled as the sole first-line therapeutic regimen administered in compartment *T*, carrying defined conditional probabilities *ω_c_* for the emergence of strain-specific resistance during active treatment (*T*^0^ → *T^c^* and *T^t^* → *T^d^*^2^). The directional selective and protective forces of the interventions modulate the effective force of infection (*λ*) via state-specific modifiers: Doxy-PEP and vaccination together reduce the effective force of infection (*U* → *E*^0^ and *U* → *E^c^*; yellow arrows in stratum *V*) by 1 − *e_vd_*. Vaccination reduces the effective force of infection uniformly across all four bacterial strains *(U* → *E*^0^, *U* → *E^c^*, *U* → *E^t^*, and *U* → *E^d^*^2^; blue arrows in strata *V* and *M*) by *e_v_*. Doxy-PEP standalone intervention acts as an ecological filter, suppressing incoming baseline susceptible and ceftriaxone-resistant exposures (*U* → *E*^0^ and *U* → *E^c^*; green arrows in stratum *D*) by *e_d_*, while leaving the transmissibility of tetracycline-resistant and dual-resistant lineages unaltered (*U* → *E^t^* and *U* → *E^d^*^2^; black arrows in strata *N* and *D*). Crucially, active doxy-PEP exposure introduces a non-zero probability for the *de novo* selection of tetracycline resistance during prophylaxis (dotted red arrows in strata *D* and *V*) with a probability *ω_t_*. Policy transitions and immunity dynamics dictate movement between the four coverage layers: doxy-PEP initiation drives flows from *N* → *D* and *M* → *V* (pink arrows) with a probability *p_d_*, while vaccine rollout shifts individuals from *N* → *M* and *D* → *V* (dark green arrows) with a probability *p_v_*. Conversely, the cessation or discontinuation of doxy- PEP reverts individuals from *D* → *N* and *V* → *M* (dark blue arrows) at a rate *ξ_n_*, whereas the temporal waning of vaccine-induced protection returns individuals from *V* → *D* and *M* → *N* (dark orange arrows) at a rate *ξ_d_*.

Although doxy-PEP exhibits high efficacy against *Treponema pallidum* (> 80%) and *Chlamydia trachomatis* ( > 70% ) [39], its efficacy against *N. gonorrhoeae* (≈ 55% ) is moderate [40]. Crucially, the real-world effectiveness of doxy-PEP against *N. gonorrhoeae* has also been found to be further compromised by the high baseline prevalence of circulating Tet-R strains [8]). Consequently, doxy-PEP selectively clears baseline- susceptible lineages while driving the clonal expansion of pre-existing or *de novo* Tet-R variants [41]. We mathematically formalize this dynamic via: (**1**) Prophylactic selection: The introduction of population-level doxy-PEP for syphilis introduces a selective force that can lead to the emergence of the Tet-R strain for *N. gonorrhoeae*; (**2**) Therapeutic selection: Continued reliance on ceftriaxone monotherapy as the first-line treatment for gonorrhoea maintains a baseline selective pressure for Cef-R emergence [42]. More importantly, tracking the intersection of doxycycline prophylaxis and ceftriaxone treatment failure provides direct epidemiological utility for public health stakeholders by mapping the change in the composition of circulating strains. Conversely, within this framework, a moderate-protection vaccine is modelled to reduce the absolute transmissibility of all circulating strains in the same manner, thereby reducing disease burden without imposing selection pressure on specific strains.

To capture the epidemiological impact of behavioural heterogeneity on transmission dynamics and intervention uptake, the model partitions the MSM population into two distinct sexual activity classes. These classes are defined by annual partner acquisition rates: high-activity individuals (≥ 5 sexual partners per annum) and low- activity individuals (< 5 sexual partners per annum). This risk-group definition was originally derived from empirical survey data [43], maintaining consistency both with established transmission modelling literature [20], [30] and the UK Health Security Agency (UKHSA) healthcare practitioner guidance for the targeted rollout of the 4CMenB vaccine [44]. The population within each behavioural group is further stratified into four intervention groups based on uptake of doxy-PEP and vaccination, representing individuals utilizing neither intervention, individuals utilizing doxy-PEP only, individuals utilizing vaccination only, individuals utilizing both doxy-PEP and vaccination. Individuals can move between these intervention groups due to waning of vaccine-induced immunity, and the elective discontinuation of doxy-PEP. Within each intersecting behavioural and intervention stratum, *N. gonorrhoeae* infections are distributed across the four modelled bacterial strains. Within each intersecting behavioural and intervention stratum, infections are distributed across the four modelled bacterial strains, each characterised by a strain-specific relative fitness parameter *f* that scales its effective transmissibility. Relative fitness represents the biological cost associated with antimicrobial resistance and determines the competitive ability of each strain in the absence of antibiotic selection. Consequently, resistant strains experience reduced transmission potential relative to the baseline susceptible strain unless sustained selective pressure from doxy-PEP or ceftriaxone treatment offsets this fitness disadvantage. The model explicitly characterizes (**1**) therapeutic treatment failure, i.e., the persistent infectivity and prolonged transmission window of individuals with Cef-R or Dual-R infections following standard-of-care ceftriaxone monotherapy, (**2**) direct selected emergence, i.e., emergence of Cef-R strains during clinical treatment of a baseline susceptible or Tet-R infection, and (**3**) bystander selected emergence and concomitant dual selection, i.e., the emergence of Tet-R or Dual-R strains driven by doxy-PEP for syphilis in an individual concurrently harbouring a baseline susceptible or Cef-R infection.

### 4. Model assumptions and proxies

*N. gonorrhoeae* can colonize multiple anatomical sites, specifically the urogenital tract, rectum, and pharynx, with each site exhibiting distinct transmission probabilities and clinical propensities for symptomatic development [45]. However, because national longitudinal surveillance data are not routinely disaggregated by anatomical site of infection, our model structural parameters represent weighted, site-averaged values across these sites. Consistent with empirical evidence, the model assumes that natural clearance or successful antibiotic treatment of a gonococcal infection does not confer protective, lasting homotypic immunity, leaving individuals immediately susceptible to reinfection upon re-exposure [46], [47]. In addition, the ceftriaxone treatment failure parameter is modelled as a static probability. While empirical treatment failure rates can vary over time and across distinct clinical settings, our static parameterized boundaries are designed to represent a uniform population-level average. Moreover, for analytical simplicity, intervention initiation is restricted to the uninfected (*U*) state. Transitions across the four intervention coverage layers (*N*, *D*, *V*, *M* ) are strictly stepwise. At any single screening event, uninfected individuals can only traverse a single arrow interface (*N* → *D*, *N* → *M*). While initial entry into an intervention stratum from the baseline pool (*N*) is mutually exclusive, individuals can transition into the dual- intervention compartment (*V*) during subsequent clinical encounters. This sequential framework assumes that joint intervention coverage is achieved via iterative clinic visits over time rather than simultaneous dual allocation at a single encounter.

A key assumption of our model is that ceftriaxone or tetracycline resistance are respectively governed by independent genetic loci. Consequently, we assume no direct genetic linkage or intrinsic biological co-selection mechanisms between these resistance determinants; any observed epidemiological co-selection is driven exclusively by ecological replacement and population-level antibiotic selection pressures. While high-resolution genomic data tracking the precise intra-strain plasmidic or chromosomal linkage of these specific determinants were not systematically available across the full calibration timeline, empirical evidence suggests that multi-drug resistance in *N. gonorrhoeae* predominantly emerges through sequential, unlinked structural mutations under distinct clinical selection pressures [48]. Furthermore, direct strain-specific surveillance data for doxycycline susceptibility were unavailable, as doxycycline minimum inhibitory concentrations (MICs) are not routinely measured within national surveillance frameworks. To address this data constraint and maintain consistency with standard public health reporting, we utilized a tetracycline MIC breakpoint of > 1 mg/L among MSM isolates as the operational proxy for doxycycline non-susceptibility [33].

### 5. Model calibration and projection

The model was parameterized and calibrated using two primary data streams: (1) Epidemiological and resistance surveillance data: National longitudinal surveillance data for MSM in England spanning 2015 to 2022 were extracted from two complementary public health registries maintained by the UKHSA. This period predates the implementation of doxy-PEP within routine clinical practice in England and therefore represents the natural epidemiological and antimicrobial resistance dynamics of *N. gonorrhoeae* in the absence of population-level doxy-PEP. Consequently, all model calibration was performed using pre-intervention surveillance data, while doxy-PEP and vaccination were introduced only during the forward projection period. Annual counts of gonorrhoea diagnoses, total diagnostic tests administered, and the corresponding proportions of Tet-R (MIC > 1mg/L) and Cef-R (MIC > 0.125mg/L) isolates were retrieved from GUMCAD [33]. Stratified annual data detailing symptomatic versus asymptomatic presentations and respective surveillance sample sizes were sourced from GRASP [34]; (2) Behavioural parameterization: Baseline behavioural inputs for the England context, specifically the structural proportions of the MSM population assigned to each sexual activity class and their respective annual partner acquisition rates, were obtained from validated empirical estimates in prior literature [20].

First, we calibrated the transmission model within a Bayesian framework to fit the empirical trajectory of annual gonorrhoea diagnoses, testing volumes, and strain-specific resistance percentages observed between 2015 and 2022 (Supplementary Information Fig. 1). Posterior distributions of the unobserved transition parameters were sampled using Hamiltonian Monte Carlo (HMC) algorithms implemented via the RStan package (version 2.32.7) in R (version 4.5.0). We executed six parallel Markov chain Monte Carlo (MCMC) chains for 2,000 iterations each, discarding the initial 1,000 iterations per chain as a burn-in period to eliminate transient initialization bias. MCMC convergence was evaluated using standard numerical and visual diagnostics, including visual inspection of sampling traceplots and analysis of marginal posterior distributions [49], monitoring of effective sample sizes (ESS) [50], and verification that the Gelman-Rubin (GR) statistic approached 1.00 for all estimated parameters [50], [51].

Next, to propagate parameter uncertainty into epidemiological projections, we extracted a random sample of 1,000 parameter combinations from the joint posterior distribution. Using the deSolve package (version 1.40), these parameter sets were used to simulate forward-looking transmission dynamics over a 15-year projection horizon from 2027 to 2041 within the MSM population in England (Fig. 1, Supplementary Information Fig. 2). We evaluated the intersection of doxy-PEP and vaccination coverage across two distinct counterfactual behaviour baselines to characterize future sexual activity and healthcare utilization: (**A**) A stabilized behaviour scenario, in which we assumed that time-varying behavioural parameters inferred during the historical calibration period, specifically the baseline force of infection and the asymptomatic screening rate, stabilized and remained constant at 2022 values throughout the projection window; (**B**) A continued trend scenario, in which we assumed that historical trajectories in sexual behaviour and screening frequency continued to evolve linearly through to 2041, extrapolating the structural trends calculated from past incidence data.

### 6. Evaluation metrics

The long-term population-level effects of the simulated interventions were quantified using two primary metrics. First, we calculated the cumulative incident infections averted/gained as well as the cumulative percentage decrease/increase for each of the four co-circulating strains (baseline susceptible, Tet-R, Cef-R, and Dual-R) over the entire 15-year projection horizon (2027-2041) relative to the counterfactual no-intervention baseline. Second, we computed the final relative strain composition at the final simulation endpoint (Year 15; 2041). This metric defines the final proportion of each distinct strain within the infected population pool. In both metrics, infections are quantified as the incidence of all newly acquired infections, which captures all individuals entering the Exposed *E* compartment, encompassing the entire cohort of hosts harbouring *N. gonorrhoeae* irrespective of symptomatic presentation or subclinical status (i.e., both symptomatic and asymptomatic infections). Moreover, to evaluate the programmatic efficiency of each intervention, we quantified the cumulative number of programme enrolments alongside the number of infections averted per enrolment over the 15-year horizon. Here, an “enrolment” is defined operationally as an eligible individual accepting the offer of doxy-PEP and/or vaccination at a sexual health clinic, rather than tracking the precise quantity of doxycycline and/or vaccine doses subsequently consumed. The no-intervention (baseline) scenario is defined as the counterfactual in which neither doxy-PEP nor vaccination is implemented throughout the simulation period. All other epidemiological, demographic, behavioural, and treatment-related parameters are held identical to the intervention scenarios. Consequently, differences between intervention and baseline projections quantify the incremental effect attributable solely to the intervention strategies rather than to underlying transmission dynamics or behavioural changes. Thresholds reported throughout the analysis are therefore derived relative to this unexposed baseline scenario.

## Results

To isolate the direct epidemiological and evolutionary impacts of doxy-PEP and vaccination from background behavioural changes, the primary analysis was evaluated based on a stabilized behavioural scenario (refer to Scenario A detailed in Methods subsection 5). Because unconstrained linear extrapolation of recent historical increases in gonorrhoea incidence may artificially inflate downstream disease forecasts, fixing behavioural parameters, specifically the force of infection and the asymptomatic screening rate at their final calibrated 2022 values provides a conservative counterfactual framework. Under this paradigm, observed shifts in transmission dynamics are attributable strictly to intervention effects rather than ongoing secular trends in risk behaviour or healthcare utilization (Fig. 2). Projections for different transmission trajectories were evaluated via sensitivity analyses and presented in the Supplementary Information.

**Fig. 2:**
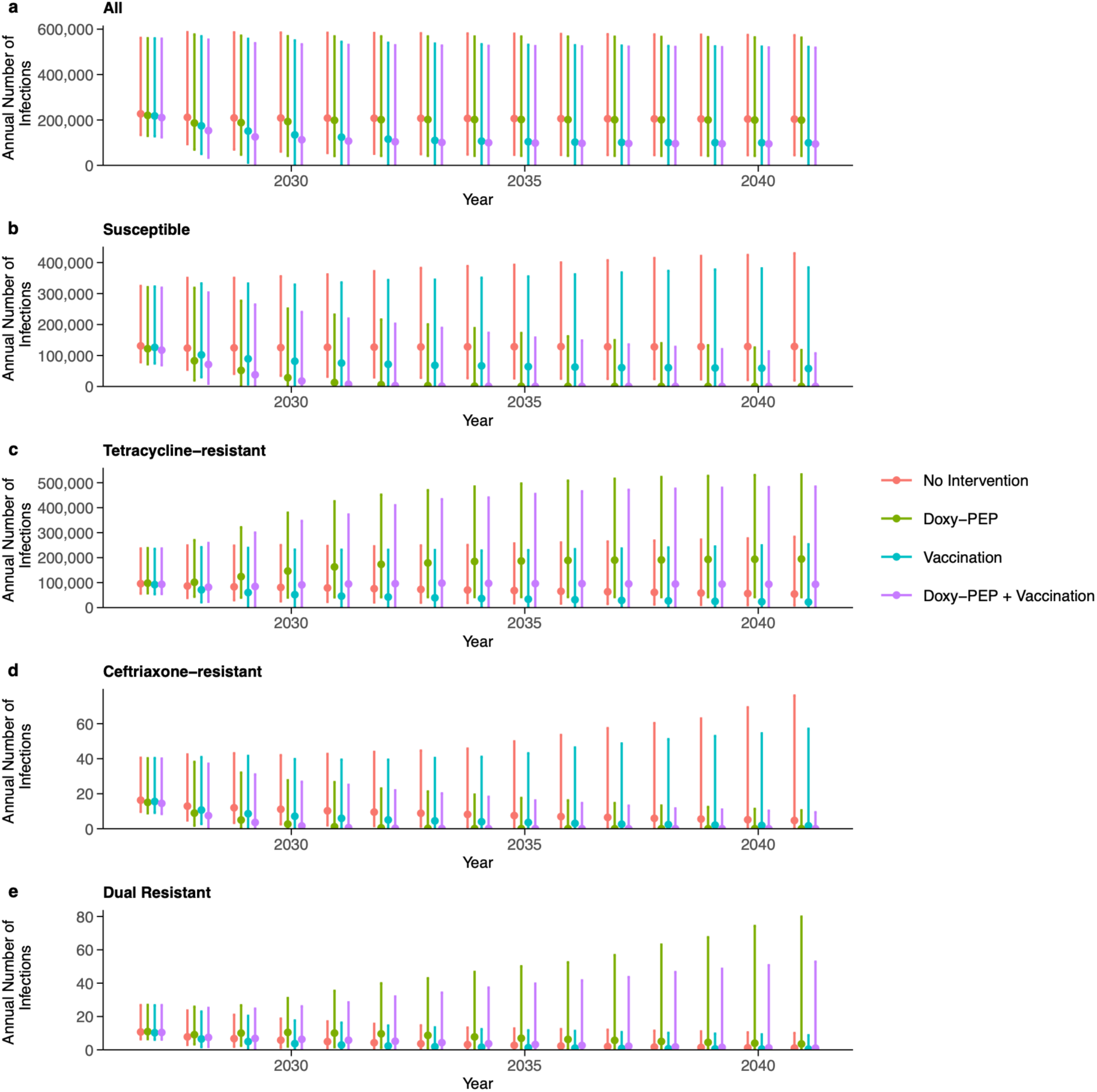
Simulated long-term impact of doxy-PEP and moderate-efficacy vaccination on *N. gonorrhoeae* incidence and resistance dynamics under baseline treatment efficacy. For each point plot, dots denote median estimates, and the accompanying vertical bars represent the corresponding 95% CrI. Projections span a 15-year horizon (2027-2041) under the stabilized behavioural baseline (Scenario A), assuming a fixed 66% intervention uptake rate within the target population. Model simulations incorporate an empirically calibrated ceftriaxone therapeutic treatment failure rate *φ* derived from historical national surveillance data. Forward projections are initialized in 2027 with population-level strain-specific resistance proportions established at 1 × 10^−4^for Cef-R and 8 × 10^−5^ for Dual-R strains. Subpanels depict trajectories for total incident infections alongside disaggregated strain-specific volumes for baseline susceptible, Tet-R, Cef-R, and Dual-R strains.

We projected future strain-specific disease incidence starting in calendar year 2027. Reflecting low-level cryptic circulation and importation, the initial population-level strain-specific resistance proportions within the infected reservoir were established at 1 × 10^−4^ for the Cef-R strain and 8 × 10^−5^ for Dual-R strain [29] (documented ceftriaxone-resistant isolates have historically been constrained to travel-associated transmission networks among heterosexual cohorts rather than establishing sustained, endemic transmission loops within the local MSM network in England [14]). Uncertainty across all metrics was propagated from a random sample of 1,000 parameter combinations from the joint posterior distribution and reported as median estimates with 95% credible intervals (CrI). Annual incident infections are rounded to the nearest 100, cumulative total averted infections and cumulative intervention enrolments are rounded to the nearest 1000.

### 1. Epidemiological burdens and strain-specific resistance dynamics

Longitudinal projections of the total *N. gonorrhoeae* burden were influenced by both prophylaxis-driven selection and vaccine-mediated suppression (Fig. 2a). Under an assumed 66% intervention uptake within the target population and an empirically calibrated, near-zero ceftriaxone therapeutic treatment failure rate *φ* derived from historical national surveillance data, doxy-PEP standalone intervention initially induces a reduction in the aggregate infection incidence. This strategy drives an initial reduction in the annual incidence of infection, reaching a minimum of 188300 (95% CrI: 47000 – 571400) infections in the third year of deployment before undergoing an epidemiological rebound. However, this prophylactic benefit is attenuated within a 5-year post-implementation window due to a rapid population-level shift toward resistant strains, with the long-term annual incidence stabilizing at around 199300 (95% CrI: 41700 – 562700) infections - a threshold comparable to the unexposed baseline of 203600 (95% CrI: 44800 – 573400) infections, thereby yielding a modest cumulative total of 108500 (95% CrI: 25700 – 284900) averted infections across the 15-year horizon (3.38% [95% CrI: 0.54% – 17.93%] cumulative decrease). Conversely, implementation of a moderate-efficacy vaccine, administered either as a standalone intervention or in a combined strategy with doxy-PEP, achieves a sustained reduction in total infections, averting a cumulative total of 959100 (95% CrI: 139400 – 254000) infections (38.88% [95% CrI: 3.20% – 83.54%] cumulative decrease) and 1055400 (95% CrI: 200300 – 2647700) infections (44.10% [95% CrI: 4.18% – 86.62%] cumulative decrease) across the 15-year horizon, respectively. These interventions establish a lower absolute transmission equilibrium that remains stable through 2041 that projects 99300 (95% CrI: 0 – 521900) infections under vaccination standalone intervention compared with 94300 (95% CrI: 0 – 518300) under the combined intervention. Crucially, over the 15-year operational horizon, the long-term annual incidence of *N. gonorrhoeae* infection under the standalone doxy-PEP strategy converged with the baseline counterfactual. Specifically, under the standalone approach, the baseline susceptible strain is replaced by the Tet-R strain. Mirroring this dynamic, the long-term epidemiological trajectories of the standalone vaccination strategy and the combined dual-intervention framework also converged.

Differences in projected strain-specific incidence highlight the selection pressures exerted by targeted antibiotic prophylaxis. Under the no-intervention baseline susceptible infections increases to 129100 (95% CrI: 19300 – 430000) by year 15. However, gonococcal infection incidence by the baseline susceptible strain rapidly declines to near-complete elimination under the introduction of either doxy-PEP standalone intervention (95% CrI: 0 – 117700 infections, Fig. 2b) or the combined strategy encompassing doxy-PEP and vaccination (0 [95% CrI: 0 – 107000] infections) by year 15, with the latter accelerating the speed at which baseline susceptible strains are eliminated from circulation (Fig. 2b). Whereas in the no-intervention and vaccination-only scenarios, Tet-R infections exhibit sustained declines, falling to 54400 (95% CrI: 8600 – 283600) infections and 21900 (95% CrI: 0 – 253600) infections by year 15, respectively (with vaccination standalone intervention averting a cumulative 316500 [95% CrI: 4500 – 1120600] infections, representing a 37.61% [95% CrI: 2.92% – 83.61%] cumulative decrease) (Fig. 2c).

Conversely, the introduction of doxy-PEP overrides the intrinsic fitness disadvantage associated to the baseline susceptible strain and triggers an expansion of the Tet-R population to 194400 (95% CrI: 41700 – 533300) infections by year 15, resulting in a negative number of −1320900 (95% CrI: −3632400 – −287300) averted Tet-R infections (-116.84% [95% CrI: −304.88% – −34.59%] cumulative decrease). Notably, the combined doxy-PEP and vaccination strategy initially suppresses Tet-R incidence below baseline levels by the second year of deployment (81700 [95% CrI: 23400 – 258700] infections vs. 86500 [38700–248700] infections at baseline). However, doxy-PEP driven selective pressure results in an increase of 93200 (95% CrI: 0 – 484500) and 54400 (95% CrI: 8600 – 283600) Tet-R infections under the doxy-PEP only and combined strategy over the non-intervention counterfactual baseline by 2041 respectively.

Despite low-level initialization to simulate emergence, Cef-R and Dual-R strains failed to achieve endemic stability, exhibiting consistent downward trajectories toward extinction across all evaluated scenarios (Fig. 2d and Fig. 2e). Sensitivity analyses demonstrated that resistance expansion is highly conditional on the value for ceftriaxone treatment failure rate: increasing the treatment failure parameter allows Cef-R and Dual-R strains to proliferate once the selection pressure successfully countervails the biological fitness penalty (detailed in the following subsection). Within the primary analysis, the combined strategy mediated the most rapid clearance of the Cef-R strain, averting a cumulative total of 90 (95% CrI: 21 – 510) infections over the 15-year horizon (72.85% [95% CrI: 28.72% – 91.54%] cumulative decrease). Conversely, the vaccination standalone intervention proved most effective at accelerating the decline of the Dual-R lineage, averting a cumulative total of 12 [95% CrI: 2 – 57] infections (28.57% [95% CrI: 2.55% – 71.09%] cumulative decrease).

### 2. Efficiency of intervention strategies

To standardize the efficiency across different implementation strategies, we computed the cumulative intervention enrolments and cumulative averted infections per enrolment over the 15-year horizon.

Under an assumed 66% intervention uptake within the target population and a near-zero ceftriaxone therapeutic treatment failure rate, a profound efficiency gap emerged between the two standalone intervention frameworks (Table 3). Standalone vaccination demonstrated a massive advantage in population-level utility, yielding a median of 0.73 (95% CrI: 0.17 – 1.63) cumulative averted infections per enrolment over the 15-year horizon. In contrast, the standalone doxy-PEP strategy demonstrated significantly lower efficiency, averting a median of just 0.072 (95% CrI: 0.018 – 0.23) cumulative infections per enrolment. This establishes that on a per-person programmatic basis, vaccination is roughly 10 times more efficient at preventing overall *N. gonorrhoeae* infections than enrolling an individual in antibiotic prophylaxis. When both interventions were deployed simultaneously, the combined strategy achieved 0.83 (95% CrI: 0.22 – 1.71) cumulative averted infections per enrolment.

**Table 1.**
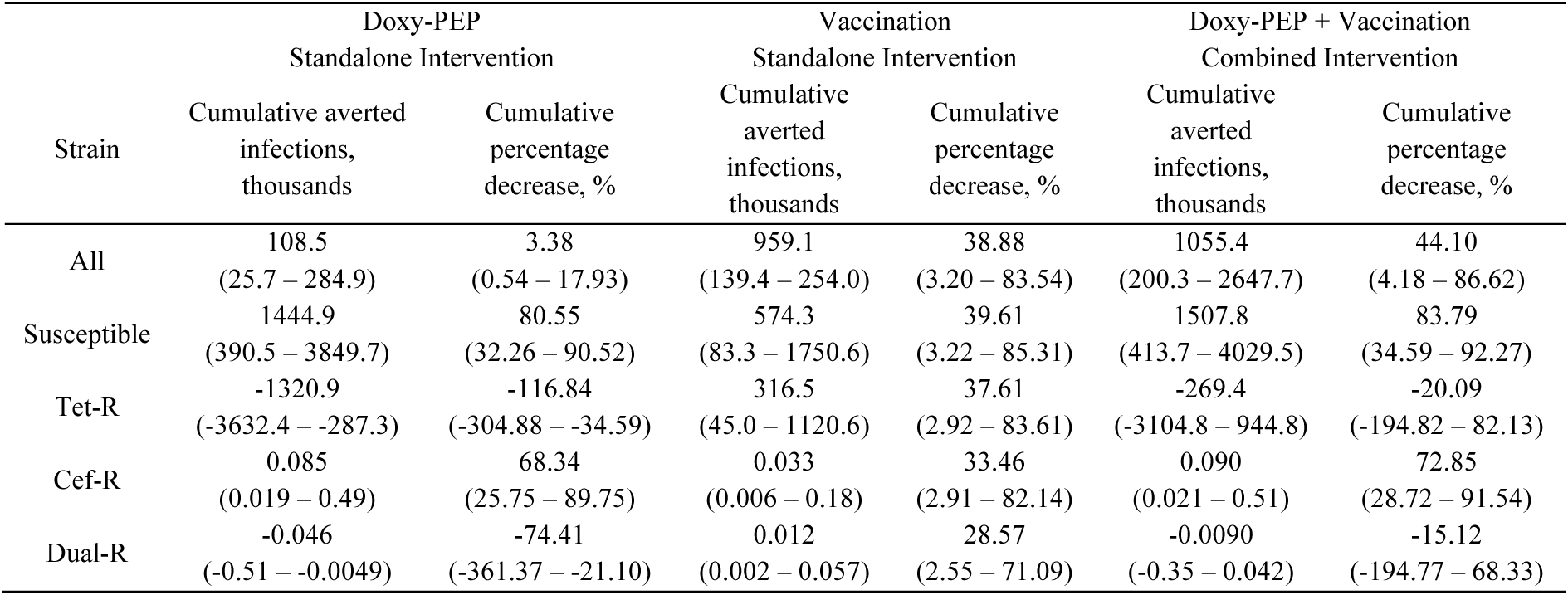
Cumulative averted infections and cumulative percentage decrease by *N. gonorrhoeae* strain and intervention strategy over a 15-year simulation horizon, 2027-2041. Cumulative averted infections are presented in thousands, and both metrics reported as median estimates with 95% CrI. Projections span from 2027 to 2041 under a stabilized behavioural baseline (Scenario A), assuming a fixed 66% intervention uptake rate within the target population. Model simulations incorporate an empirically calibrated, near-zero ceftriaxone therapeutic treatment failure rate *φ* derived from historical national surveillance data. Forward projections are initialized in 2027 with population-level strain-specific resistance proportions set at 1 × 10^−4^ for Cef-R and 8 × 10^−5^ for Dual-R strains.

**Table 3.** Cumulative intervention enrolments and cumulative averted infections per enrolment by *N. gonorrhoeae* strain and intervention strategy over a 15-year simulation horizon, 2027-2041. Cumulative intervention enrolments are presented in thousands, and both metrics reported as median estimates with 95% CrI. Projections span from 2027 to 2041 under a stabilized behavioural baseline (Scenario A), assuming a fixed 66% intervention uptake rate within the target population. Model simulations incorporate an empirically calibrated, near-zero ceftriaxone therapeutic treatment failure rate *φ* derived from historical national surveillance data. Forward projections are initialized in 2027 with population-level strain-specific resistance proportions set at 1 × 10^−4^ for Cef-R and 8 × 10^−5^ for Dual-R strains.

| Strain | Doxy-PEP<br>Standalone Intervention |  | Vaccination<br>Standalone Intervention |  | Doxy-PEP + Vaccination<br>Combined Intervention |  |
| --- | --- | --- | --- | --- | --- | --- |
|  | Cumulative doxy-PEP enrolments,<br>thousands | Cumulative<br>averted<br>infections per<br>enrolment | Cumulative<br>vaccination<br>enrolments,<br>thousands | Cumulative<br>averted<br>infections per<br>enrolment | Cumulative<br>intervention<br>enrolments,<br>thousands | Cumulative<br>averted<br>infections per<br>enrolment |
| All | 1501.7<br>(637.5 – 2729.9) | 0.072<br>(0.018 – 0.23) | 1295.1<br>(595.1 – 2070.1) | 0.73<br>(0.17 – 1.63) | 2842.8<br>(1243.4 – 4819.6) | 0.83<br>(0.22 – 1.71) |
| Susceptible | - | 0.99<br>(0.18 – 3.69) | - | 0.45<br>(0.096 – 1.09) | - | 1.20<br>(0.24 – 4.20) |
| Tet-R | - | -0.93<br>(-3.46 – -0.13) | - | 0.24<br>(0.054 – 0.75) | - | -0.23<br>(-3.37 – 0.59) |
| Cef-R | - | 0<br>(0 – 0.00051) | - | 0<br>(0 – 0.00013) | - | 0<br>(0 – 0.00059) |
| Dual-R | - | 0<br>(-0.00039 – 0) | - | 0<br>(0 – 0) | - | 0<br>(-0.00037 – 0) |

Disaggregating these metrics by individual resistance profiles in Table 3 uncovers the underlying evolutionary trade-offs and selective pressures driving these efficiency profiles. While standalone doxy-PEP averts baseline susceptible strains by 0.99 (95% CrI: 0.18 – 3.69) cumulative infections per enrolment, it generates a substantial net expansion for Tet-R strains, averting −0.93 (95% CrI: −3.46 – −0.13) cumulative infections per enrolment. This explicitly quantifies the population-level speed of competitive release, demonstrating that for approximately every single individual enrolled in a doxy-PEP regimen, the program structurally shifts transmission dynamics to produce nearly one additional Tet-R case. Conversely, standalone vaccination maintains an evolutionary-neutral profile, averting 0.45 (95% CrI: 0.096 – 1.09) baseline susceptible infections and 0.24 (95% CrI: 0.054 – 0.75) Tet-R infections per enrolment without driving net negative infections for any circulating lineage. Crucially, under the dual intervention strategy, the addition of the vaccine significantly suppresses the evolutionary penalty of prophylaxis. The combined approach achieves a median of 1.20 (95% CrI: 0.24 – 4.20) averted baseline susceptible infections, while dramatically blunting the Tet-R expansion down to a less severe efficiency of −0.23 (95% CrI: −3.37 – −0.59) averted infections per enrolment.

### 3. Impact of ceftriaxone treatment failure on strain dynamics and programmatic efficiency

To evaluate the impact of treatment failure on strain dynamics, under an assumed 66% intervention uptake within the target population, sensitivity analysis was conducted by increasing the ceftriaxone therapeutic treatment failure parameter *φ* to 20%. This sensitivity analysis simulates a large increase in treatment failure due to ceftriaxone over empirically observed rates (Fig. 3). Under this high-resistance scenario, the epidemic trajectories for total incident infections, baseline susceptible lineages, and Tet-R strains remain consistent with that of primary analysis (Fig. 3a, Fig. 3b, and Fig.3c).

**Fig. 3:**
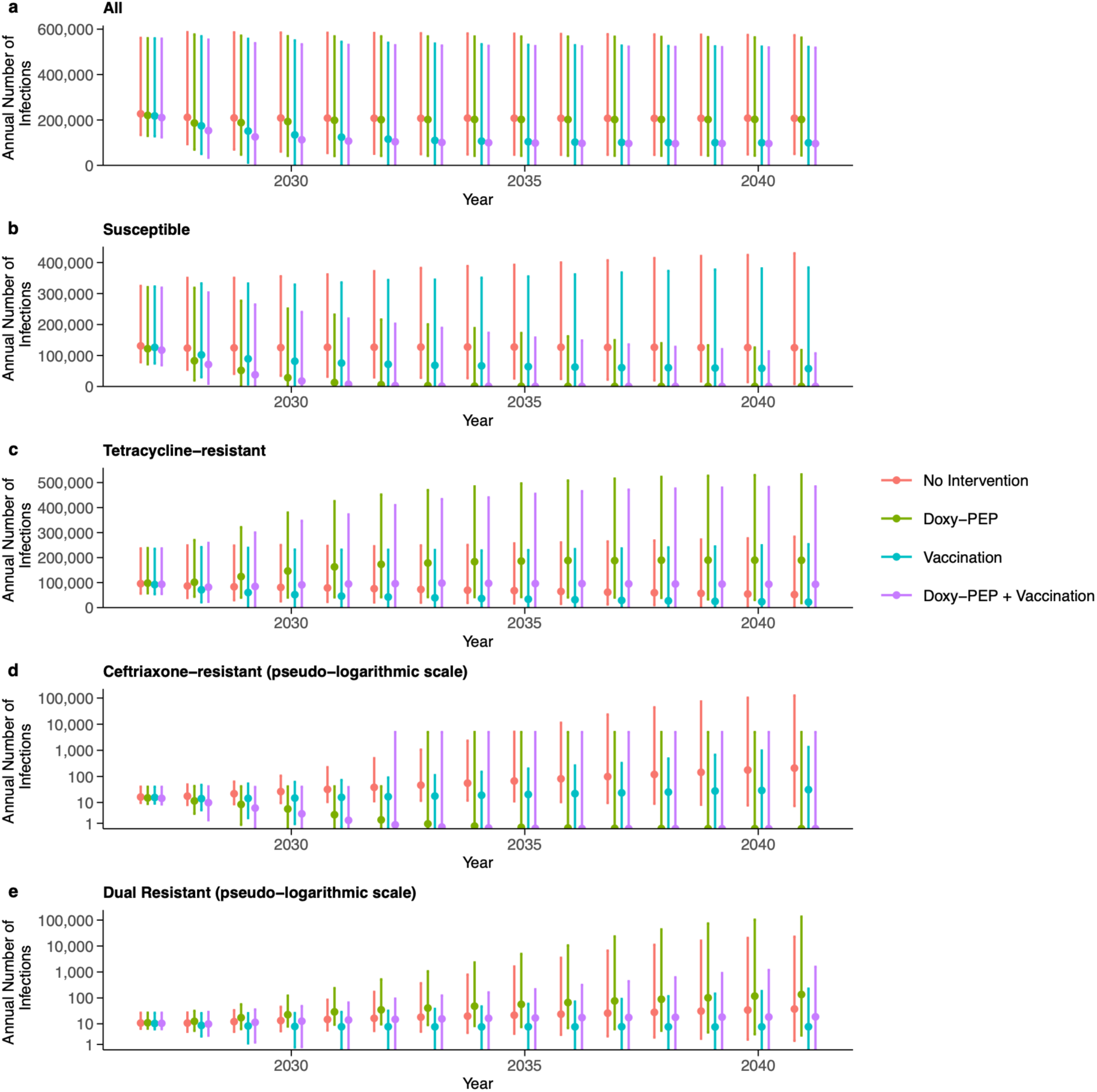
Simulated long-term impact of doxy-PEP and moderate-efficacy vaccination on *N. gonorrhoeae* transmission volumes and resistance dynamics under accelerated frontline treatment failure. For each point plot, dots denote median estimates, and the accompanying vertical error bars represent the corresponding 95% CrI. Projections span a 15-year horizon (2027-2041) under the stabilized behavioural baseline (Scenario A), assuming a fixed 66% intervention uptake rate within the target population. Model simulations incorporate an artificially elevated ceftriaxone therapeutic treatment failure rate (*φ* = 20%) to evaluate system robustness against frontline ceftriaxone compromise. Forward projections are initialized in 2027 with population-level strain- specific resistance proportions established at 1 × 10^−4^for Cef-R and 8 × 10^−5^for Dual-R strains. Subpanels depict trajectories for total incident infections alongside disaggregated strain-specific volumes for baseline susceptible, Tet-R, Cef-R, and Dual-R strains. To improve visualization across several orders of magnitude, the y-axes in panels d (Cef-R) and e (Dual-R) are displayed using a pseudo-logarithmic (*log*_10_-equivalent) scale, whereas panels a-c use a linear scale.

The most notable deviation from the main analysis (Results subsection 1) is observed in the trajectories of the Cef-R and Dual-R strains (Fig. 3d and Fig. 3e), which no longer follow a path toward extinction. At a 20% treatment failure rate, in the unexposed no-intervention scenario, there is a sustained, linear increase with significant outbreak potential, reaching 200 (95% CrI: 7 – 125100) annual incident infections by year 15. This is in contrast to the downward trend observed at the main analysis (Results subsection 1). Crucially, while the doxy- PEP standalone strategy, vaccination standalone strategy, and the combined dual-intervention strategy all remain effective at suppressing the standalone Cef-R infections to near-zero levels, averting a cumulative total of 1000 (95% CrI: 100 – 390900) infections (94.09% [95% CrI: 32.49% – 99.99%] cumulative decrease), 370 (95% CrI: 13 – 390900) infections (45.19% [95% CrI: 3.34% – 99.98%] cumulative decrease), and 1000 (95% CrI: 110 – 390900) infections (95.42% [95% CrI: 35.13% – 100.00%] cumulative decrease), respectively. Notably, the introduction of targeted antibiotic prophylaxis leads to a clear shift in the trajectories of Dual-R strain infections. Under the doxy-PEP standalone intervention, Dual-R incidence increases to 136 (95% CrI: 3 – 135700) infections by year 15, exceeding the no-intervention scenario of 37 (95% CrI: 2 – 22900) infections by the 2041 horizon, yielding a negative cumulative total of -510 (95% CrI: −258600 – −38) averted infections across the 15-year horizon (-146.18% [95% CrI: -920.86% – −31.30%] cumulative decrease).

In contrast, vaccination alone and the combined strategy mitigates AMR emergence, successfully suppressing both Cef-R and Dual-R lineages. For the Cef-R strain, the vaccination standalone strategy averts a cumulative total of 370 (95% CrI: 13 - 390900) infections (45.19% [95% CrI: 3.34% – 99.98%] cumulative decrease), while the combined strategy averts 100 (95% CrI: 5 – 90100) total infections (95.42% [95% CrI: 35.13% – 100.00%] cumulative decrease). For the Dual-R strain, the vaccination standalone strategy averts a cumulative total of 1000 (95% CrI: 110 – 390900) infections (40.79% [95% CrI: 3.06% – 99.94%] cumulative decrease), whereas the combined strategy yields -41 (95% CrI: −3100 – 86200) infections (-24.41% [95% CrI: −387.51% – 99.91%] cumulative decrease). By year 15, Cef-R annual incidence falls to 31 (95% CrI: 0 – 1300) infections under vaccination standalone strategy compared with 0 (95% CrI: 0 – 38) infections under the combined strategy. Similarly, Dual-R annual incidence is restricted to 8 (95% CrI: 0 – 230) infections under vaccination compared with 19 (95% CrI: 0 – 1600) infection under the dual-intervention approach.

Moreover, for the efficiency of different intervention strategies, under this high treatment failure scenario, the cumulative intervention enrolments and the net aggregate efficiencies across all intervention strategies (Table 5) remain stable (as compared to Table 3), but changes the underlying strain-specific composition. For standalone doxy-PEP, the per-enrolment cumulative averted infections for Dual-R strains drops below zero into negative territory, reaching −0.00039 (95% CrI: −0.12 – 0). In contrast, the dual intervention strategy holds the Dual-R per- enrolment cumulative averted infections flat at 0 (95% CrI: −0.0029 – 0.042).

**Table 4.** Cumulative averted infections and cumulative percentage decrease by *N. gonorrhoeae* strain and intervention strategy over a 15-year simulation horizon, 2027-2041. Cumulative averted infections are presented in thousands, and both metrics reported as median estimates with 95% CrI. Projections span from 2027 to 2041 under a stabilized behavioural baseline (Scenario A), assuming a fixed 66% intervention uptake rate within the target population. Model simulations incorporate an elevated ceftriaxone therapeutic treatment failure rate (*φ* = 20%). Forward projections are initialized in 2027 with population-level strain-specific resistance proportions set at 1 × 10^−4^ for Cef-R and 8 × 10^−5^ for Dual-R strains.

| r | Doxy-PEP<br>Standalone Intervention |  | Vaccination<br>Standalone Intervention |  | Doxy-PEP + Vaccination<br>Combined Intervention |  |
| --- | --- | --- | --- | --- | --- | --- |
|  | Cumulative<br>averted infections,<br>thousands | Cumulative<br>percentage<br>decrease, % | Cumulative<br>averted<br>infections,<br>thousands | Cumulative<br>percentage<br>decrease, % | Cumulative<br>averted<br>infections,<br>thousands | Cumulative<br>percentage<br>decrease, % |
| All | 110.1<br>(25.7 – 296.3) | 3.43<br>(0.54 – 17.78) | 964.9<br>(139.4 – 256.1) | 38.87<br>(3.20 – 84.23) | 1069.0<br>(200.3 – 2678.2) | 44.09<br>(4.18 – 87.03) |
| Susceptible | 1420.4<br>(385.0 – 3849.5) | 80.25<br>(32.26 – 90.13) | 559.3<br>(83.2 – 1619.0) | 39.59<br>(3.22 – 84.46) | 1484.8<br>(408.7 – 4028.6) | 83.64<br>(34.59 – 92.05) |
| Tet-R | -1310.2<br>(-3630.7 – -285.9) | -116.91<br>(-297.66 – -34.60) | 310.4<br>(45.0 – 1164.8) | 37.61<br>(2.92 – 82.86) | -269.3<br>(-3101.2 – 926.4) | -20.09<br>(-194.62 – 81.02) |
| Cef-R | 1.0<br>(0.10 – 390.9) | 94.09<br>(32.49 – 99.99) | 0.37<br>(0.013 – 390.9) | 45.19<br>(3.34 – 99.98) | 1.0<br>(0.11 – 390.9) | 95.42<br>(35.13 – 100.00) |
| Dual-R | -0.51<br>(-258.6 – -0.038) | -146.18<br>(-920.86 – -31.30) | 0.10<br>(0.005 – 90.1) | 40.79<br>(3.06 – 99.94) | -0.041<br>(-3.1 – 86.2) | -24.41<br>(-387.51 – 99.91) |

**Table 5.** Cumulative intervention enrolments and cumulative averted infections per enrolment by *N. gonorrhoeae* strain and intervention strategy over a 15-year simulation horizon, 2027-2041. Cumulative intervention enrolments are presented in thousands, and both metrics reported as median estimates with 95% CrI. Projections span from 2027 to 2041 under a stabilized behavioural baseline (Scenario A), assuming a fixed 66% intervention uptake rate within the target population. Model simulations incorporate an elevated ceftriaxone therapeutic treatment failure rate (*φ* = 20%). Forward projections are initialized in 2027 with population-level strain-specific resistance proportions set at 1 × 10^−4^ for Cef-R and 8 × 10^−5^ for Dual-R strains.

| Strain | Doxy-PEP<br>Standalone Intervention |  | Vaccination<br>Standalone Intervention |  | Doxy-PEP + Vaccination<br>Combined Intervention |  |
| --- | --- | --- | --- | --- | --- | --- |
|  | Cumulative doxy-PEP enrolments,<br>thousands | Cumulative<br>averted<br>infections per<br>enrolment | Cumulative<br>vaccination<br>enrolments,<br>thousands | Cumulative<br>averted<br>infections per<br>enrolment | Cumulative<br>intervention<br>enrolments,<br>thousands | Cumulative<br>averted infections<br>per enrolment |
| All | 1501.6<br>(637.5 – 2728.0) | 0.073<br>(0.018 – 0.23) | 1295.1<br>(595.1 – 2070.1) | 0.73<br>(0.18 – 1.64) | 2842.8<br>(1243.3 – 4819.6) | 0.84<br>(0.22 – 1.79) |
| Susceptible | - | 0.99<br>(0.18 – 3.68) | - | 0.44<br>(0.096 – 1.05) | - | 1.20<br>(0.24 – 4.20) |
| Tet-R | - | -0.91<br>(-3.46 – -0.12) | - | 0.23<br>(0.054 – 0.74) | - | -0.23<br>(-3.37 – 0.56) |
| Cef-R | - | 0.00074<br>(0 – 0.15) | - | 0.00030<br>(0 – 0.20) | - | 0.00087<br>(0.00011 – 0.20) |
| Dual-R | - | -0.00039<br>(-0.12 – 0) | - | 0<br>(0 – 0.042) | - | 0<br>(-0.0029 – 0.042) |

### 4. Sensitivity analyses under varied ceftriaxone treatment failure rate and intervention uptake

Next, to distinguish the relative impact of ceftriaxone treatment failure rates and intervention uptake on long-term epidemiological outcomes, we conducted a sensitivity analysis to determine whether the frontline therapeutic treatment failure rate or absolute intervention uptake rates serve as the primary driver of final strain dominance. Specifically, we quantified the final relative strain composition (%) of baseline susceptible, Tet-R, Cef-R, and Dual-R infections at the 15-year simulation horizon (2041). These results reflect median projections generated from a probabilistic sensitivity analysis sampling 1000 parameter combinations from the joint posterior distribution. Moreover, these outcomes were analysed by jointly varying the ceftriaxone treatment failure rate (ranging from 0.0 to 1.0) and target population uptake rates (ranging from 0.0 to 1.0) for no intervention, doxy- PEP standalone intervention, vaccination standalone intervention, and the combined intervention strategy (see Fig. 4A).

**Fig. 4:**
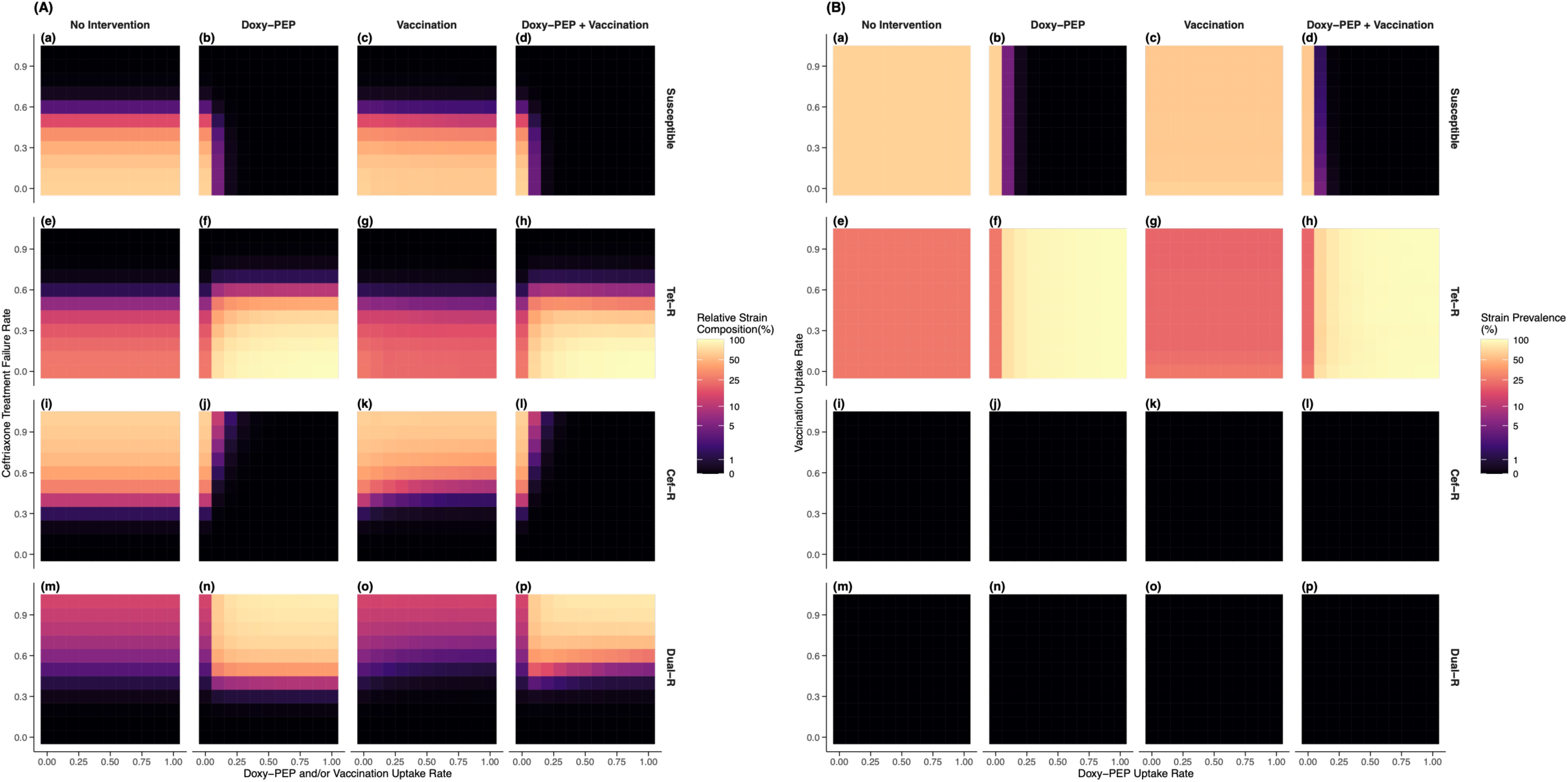
Long-term strain composition under varying ceftriaxone treatment failure and intervention uptake. **Panel A.** Predicted relative strain composition at the 15-year simulation horizon (2041) across varying ceftriaxone treatment failure rates and intervention strategies. Each heatmap cell represents the median projection from a probabilistic sensitivity analysis based on 1000 parameter sets sampled from the joint posterior distribution. Simulations assume the stabilised behavioural scenario (Scenario A) and initial 2027 resistance frequencies of 1 × 10^−4^for Cef-R and 8 × 10^−5^ for Dual-R strains. Columns correspond to the four intervention strategies (no intervention, doxy-PEP alone, vaccination alone, and combined doxy-PEP plus vaccination), while rows represent the four circulating strains (susceptible, Tet-R, Cef-R, and Dual-R). Within each heatmap, the horizontal axis denotes intervention uptake (0-100%) and the vertical axis denotes the frontline ceftriaxone treatment failure rate (0-100%). Colours indicate the proportion of all active gonococcal infections attributable to each strain at year 2041. Values are displayed using a pseudo-logarithmic (*log*_10_-equivalent) colour scale with thresholds of 0%, 1%, 5%, 10%, 25%, 50%, and 100% to distinguish early competitive shifts from complete strain replacement. **Panel B.** Predicted relative strain composition at the 15-year simulation horizon (2041) across joint intervention uptake levels. Simulations use the same posterior parameter samples, behavioural assumptions, and initial resistance frequencies as Panel A. Columns again correspond to the four intervention strategies, whereas rows correspond to the four circulating strains. Within each heatmap, the horizontal axis represents doxy-PEP uptake (0-100%) and the vertical axis represents vaccination uptake (0-100%). Colours indicate the proportion of all active gonococcal infections attributable to each strain at year 2041 using the same pseudo-logarithmic colour scale as Panel A.

Under the no-intervention baseline and vaccination standalone intervention scenarios, strain distribution is governed primarily by treatment efficacy. At low treatment failure rates, the reservoir remains overwhelmingly dominated by the baseline susceptible strain (Fig. 4Aa and Fig. 4Ac). Conversely, as frontline treatment failure scales toward a complete loss of ceftriaxone efficacy, the baseline susceptible population is eradicated and replaced by the Cef-R strain (Fig. 4Ai and Fig. 4Ak). A fundamental ecological shift occurs upon the introduction of doxy-PEP standalone intervention or the combined strategy. Across all intervention scenarios, low ceftriaxone treatment failure rates select heavily for the Tet-R strain (Fig. 4Af and Fig. 4Ah). However, when high antibiotic prophylaxis rates intersect with elevated ceftriaxone treatment failure, final infection incidence is dominated by the Dual-R strain (Fig. 4An and Fig. 4Ap). Within a fixed ceftriaxone failure regime, incremental increases in doxy-PEP uptake amplify the absolute selective advantage of the Tet-R variant, increasing population-level strain composition (Fig. 4Af and Fig. 4Ah).

### 5. Sensitivity analyses under varied intervention uptake of doxy-PEP and vaccination

To compare the impact of intervention-specific coverages on long-term ecological composition, we evaluated the joint effects of doxy-PEP and vaccination uptake rates under the baseline clinical scenario of an empirically calibrated, near-zero ceftriaxone therapeutic treatment failure rate. The final strain composition was quantified at the 15-year simulation horizon (2041) as the proportion of all active N. gonorrhoeae infections attributable to the baseline susceptible, Tet-R, Cef-R, and Dual-R strains. These results reflect median projections generated from a probabilistic sensitivity analysis sampling 1000 parameter combinations from the joint posterior distribution. Moreover, these outcomes were derived via continuously varying both the target population uptake rates for doxy- PEP (ranging from 0.0 to 1.0) against target population uptake rates for vaccination (ranging from 0.0 to 1.0) across all four intervention strategies (Fig. 4B).

Under both the no-intervention baseline and vaccination standalone intervention regimes, expanding vaccine coverage exerts a negligible effect on final epidemiological outcomes. In these scenarios, the population remains heavily dominated by the baseline susceptible lineage (Fig. 4Ba and Fig. 4Bc) followed by a stable, low relative strain composition of Tet-R infections (Fig. 4Be and Fig. 4Bg). Meanwhile, the absence of frontline selection pressure (near-zero ceftriaxone treatment failure rate) maintains both the Cef-R and Dual-R strains at zero (Fig. 4Bi, Fig. 4Bm, Fig. 4Bk, and Fig. 4Bo). Conversely, the introduction of doxy-PEP standalone intervention induces a shift in strain composition, dependent on coverage (Fig. 4Bb and Fig. 4Bf). As doxy-PEP uptake scales, the baseline susceptible-strain population falls to near-elimination (Fig. 4Bb), with a concurrent rapid increase in the proportion of Tet-R strains over time (Fig. 4Bf). Cef-R and Dual-R lineages remain structurally suppressed at near-zero levels (Fig. 4Bj and Fig. 4Bn). Crucially, within the combined dual-intervention strategy, the simultaneous introduction of vaccination fails to significantly alter the strain distribution observed under doxy- PEP standalone intervention (Fig. 4Bd and Fig. 4Bh).

## Discussion

This modelling study provides a quantitative evaluation of the long-term epidemiological consequences of deploying doxy-PEP for syphilis prevention and a moderately effective transmission-blocking vaccine against *N. gonorrhoeae* in MSM in England. Our projections suggest that although standalone doxy-PEP initially reduces overall gonorrhoea incidence, its epidemiological benefits progressively diminish over time owing to the expansion of Tet-R strains (Fig. 2a and Fig. 3a). Consequently, doxy-PEP alone may be insufficient to sustain long-term reductions in gonorrhoea incidence, despite its well-established effectiveness against syphilis and chlamydia [39], [40], [52], [53]. By contrast, vaccine-inclusive strategies consistently produced larger and more durable reductions in transmission because they reduce host susceptibility without imposing antibiotic selection pressure (Fig. 2 and Fig. 3). Moreover, in the absence of sustained population-level doxycycline exposure, the intrinsic fitness cost associated with Tet-R determinants renders these strains less competitive than susceptible lineages (Fig. 2b, Fig. 2c, Fig. 3b and Fig. 3c), demonstrating that increasing transmission alone is insufficient to maintain resistant strains without continued selective pressure. Under scenarios of elevated ceftriaxone treatment failure, however, widespread doxy-PEP deployment favours expansion of Dual-R strains, whereas vaccination, either alone or combined with doxy-PEP, continues to suppress overall transmission (Fig. 3e). Collectively, these findings suggest that the long-term public health utility of doxy-PEP is critically dependent on preserving the effectiveness of frontline ceftriaxone therapy, while vaccines offer an evolutionarily neutral approach to reducing gonorrhoea transmission. More importantly, across all intervention scenarios, long-term epidemiological behaviour is characterised by competitive strain replacement rather than uniform expansion of all strains. Doxy- PEP preferentially removes susceptible strains, allowing Tet-R lineages to expand through ecological release, whereas vaccine-inclusive strategies reduce transmission across all strains without altering their competitive hierarchy. Under elevated ceftriaxone treatment failure, this ecological shift further facilitated the emergence of Dual-R strains.

Our findings should be interpreted alongside emerging real-world evidence on antimicrobial resistance following doxy-PEP implementation. A recent observational study from Milan reported that approximately 20% of gonorrhoea episodes occurred among doxy-PEP users. However, baseline tetracycline resistance was already extremely high (96%), and doxy-PEP exposure was not associated with significantly increased high-level tetracycline resistance after adjustment for multiple comparisons [12]. These findings suggest that when tetracycline resistance is already close to fixation within the circulating gonococcal population, doxy-PEP may exert comparatively little additional selective pressure because susceptible strains have largely disappeared. By contrast, our model was parameterised using surveillance data from England, where tetracycline susceptibility remains substantially more common. Under these conditions, doxy-PEP preferentially suppresses susceptible strains, creating ecological space for competitive replacement by Tet-R lineages. Consequently, the magnitude of resistance selection projected in our study should be interpreted as epidemiological context-specific rather than universally applicable and will depend on the baseline prevalence of tetracycline resistance in the target population.

Although vaccine-inclusive strategies consistently outperformed doxy-PEP alone in our simulations, it is important to emphasise that such vaccination strategies remain hypothetical for gonorrhoea prevention at present. While observational studies have suggested cross-protection of the outer membrane vesicle meningococcal B vaccines against gonorrhoea, recent randomised evidence has been less encouraging. The DOXYVAC trial reported a non-significant reduction in gonorrhoea incidence following 4CMenB vaccination [39], and preliminary findings from the GoGoVax trial likewise failed to demonstrate statistically significant protection [22]. These findings highlight the biological challenge of developing an effective gonococcal vaccine, particularly given that natural gonococcal infection itself induces little durable protective immunity. Accordingly, our analyses should be interpreted as evaluating the potential population-level value of a moderately effective transmission-blocking vaccine rather than predicting the effectiveness of any currently licensed vaccination programme. Nevertheless, our results consistently demonstrate that interventions capable of reducing transmission without imposing antimicrobial selection pressure provide the greatest long-term protection against antimicrobial resistance.

Sensitivity analyses further demonstrated that although intervention uptake substantially influenced the magnitude of transmission reduction and the rate of strain replacement, the eventual long-term strain composition was governed primarily by the background ceftriaxone treatment failure rate rather than by intervention coverage itself (Fig. 2 and Fig. 3). Likewise, varying vaccine uptake produced relatively little effect on equilibrium strain composition (Fig. 4B), indicating that the selective pressure generated by doxy-PEP dominates long-term resistance dynamics once antibiotic exposure becomes widespread. These findings emphasise that preserving the clinical effectiveness of ceftriaxone is likely to be more influential for long-term antimicrobial resistance containment than modest differences in intervention uptake alone.

Our framework substantially extends previous mathematical modelling studies evaluating doxy-PEP and gonococcal vaccination. Existing models have largely examined these interventions independently, characterising either the public health impact of vaccination in isolation [20], [30], [31] or the epidemiological benefits of doxy- PEP for syphilis prevention [2]. More recent gonorrhoea models have begun to investigate the selective pressures exerted by doxy-PEP on antimicrobial resistance, but these analyses have primarily been restricted to single-strain transmission dynamics [8] or standalone intervention scenarios [12], [29]. In contrast, our model explicitly captures the concurrent ecological and evolutionary interactions among four co-circulating gonococcal strains under simultaneous intervention pressures. By integrating susceptible, tetracycline-resistant, ceftriaxone-resistant, and dual-resistant strains within a unified transmission framework, our analyses demonstrate how an intervention directed at one pathogen or resistance mechanism can unintentionally reshape the epidemiology of another. These findings underscore the importance of evaluating preventive interventions within a broader ecological context that explicitly considers antimicrobial selection and strain replacement rather than focusing solely on short-term reductions in disease incidence.

Several limitations should be considered when interpreting these findings. First, the model was calibrated using surveillance data from England and therefore reflects the epidemiological characteristics and antimicrobial resistance landscape of a single country. Although England provides comprehensive longitudinal surveillance suitable for robust model calibration, caution is warranted when extrapolating these findings to settings with substantially different resistance profiles. In particular, ceftriaxone resistance remains uncommon in England but is considerably more prevalent in several countries within the WHO Western Pacific Region, including Cambodia, China, and Japan [54]. Under such epidemiological conditions, resistant strains may follow different evolutionary trajectories, and the long-term effectiveness of doxy-PEP and vaccination may differ quantitatively. Future studies should therefore recalibrate this modelling framework using local surveillance and antimicrobial resistance data to support region-specific policy evaluation.

Second, although behavioural heterogeneity was incorporated through stratification by annual partner acquisition rates, the model does not explicitly represent heterogeneity in anatomical sites of infection or sexual practices because detailed empirical data to parameterise these processes are currently unavailable. Increasing evidence suggests that oropharyngeal transmission contributes disproportionately to gonorrhoea transmission among MSM [55], and ceftriaxone-resistant infections at different anatomical sites may differ in both transmissibility and treatment outcomes [14]. Incorporating anatomical-site-specific transmission pathways, screening practices, and treatment responses would improve biological realism and may further refine projections of antimicrobial resistance emergence and persistence as relevant epidemiological data become available.

Third, although the relative fitness parameters of the resistant strains were estimated through Bayesian calibration rather than fixed a priori, empirical data directly informing these quantities remain limited. Consequently, prior distributions for the strain-specific fitness parameters were specified from the available literature and biological plausibility, with posterior estimates inferred jointly with the remaining model parameters during calibration. As with all inverse modelling approaches, the inferred fitness values are therefore constrained by both the available surveillance data and the assumed model structure, rather than representing direct biological measurements. Because relative fitness governs the competitive dynamics between susceptible and resistant lineages, uncertainty in these posterior estimates remains an important source of uncertainty in the projected long-term resistance trajectories, particularly during the early stages of antimicrobial resistance emergence. Future experimental and epidemiological studies that directly quantify the transmission fitness of Tet-R, Cef-R, and Dual-R *N. gonorrhoeae* strains will further improve the precision of these projections.

Lastly, our modelling framework incorporates two structural assumptions that, if anything, are likely to render our projections conservative. Transitions between intervention strata were assumed to occur sequentially. Although simultaneous initiation of doxy-PEP and vaccination is clinically feasible during a single healthcare encounter, the sequential structure provides a pragmatic representation of population-level implementation processes, including staggered programme roll-out, appointment scheduling, and delays in intervention uptake. Consequently, the model may modestly underestimate the speed at which individuals achieve dual-intervention coverage and therefore underestimate the short-term effectiveness of combined intervention strategies. Second, model parameterisation was informed primarily by data on low-level tetracycline resistance. The emergence and persistence of high-level doxycycline resistance may be constrained by additional biological and fitness costs, although the magnitude of these costs remains uncertain. As a result, the late-stage resurgence of resistant infections observed in doxy-PEP-inclusive scenarios should be interpreted as a conservative upper-bound scenario rather than a precise prediction of future resistance trajectories. This approach intentionally errs on the side of caution by avoiding optimistic assumptions regarding resistance dynamics. Importantly, these two assumptions influence projections in opposite directions: the sequential uptake assumption may underestimate intervention effectiveness, whereas the resistance assumptions may overestimate the long-term burden of resistance. Taken together, they establish a deliberately conservative framework that minimises the risk of overstating the long-term public health benefits of either intervention.

Future research should first explicitly quantify the long-term clinical and economic healthcare burdens induced by widespread doxy-PEP implementation. Specifically, models must capture how targeted prophylaxis constructs direct evolutionary conduits that facilitate a rapid transition from standalone tetracycline resistance to multidrug-resistant strains via ceftriaxone co-selection using genomic data. Second, this cross-pathogen analytical framework should be extended to evaluate the collateral selective pressures of sustained, high-volume tetracycline exposure on other critical components of the sexual health microbiome. Incorporating the resistance kinetics of co-circulating pathogens such as *C. trachomatis* and *T. pallidum* is essential to achieving a systems-biology approach to long-term antimicrobial stewardship. Finally, future iterations must transition from single-pathogen dynamics toward an integrated, multi-STI syndromic framework. By simultaneously modelling the overlapping transmission networks, incident volumes, and clinical sequelae of *C. trachomatis*, *T. pallidum*, and *N. gonorrhoeae*, future analyses can provide holistic net utility assessments of doxy-PEP. Such evaluations are vital to ensuring that localized reductions in one bacterial infection do not inadvertently accelerate multidrug-resistant selection.

Overall, our findings suggest that although doxy-PEP provides public health benefits for preventing syphilis and chlamydia, widespread implementation may accelerate the emergence and expansion of multidrug-resistant *N. gonorrhoeae* under epidemiological conditions where tetracycline susceptibility remains common and frontline ceftriaxone effectiveness begins to decline. Conversely, interventions that reduce transmission without imposing antimicrobial selection pressure, such as effective vaccination, offer a more evolutionarily sustainable strategy for long-term gonorrhoea control. Importantly, our results should be interpreted as conditional on both the underlying antimicrobial resistance landscape and the effectiveness of future gonococcal vaccines. Together with robust antimicrobial resistance surveillance and antimicrobial stewardship, these findings support the integration of non-antibiotic preventive interventions [20], [56] into future gonorrhoea control programmes while emphasising the continued importance of preserving frontline ceftriaxone efficacy. Beyond gonorrhoea, this work illustrates a broader principle for antimicrobial stewardship: interventions designed to prevent one STI may generate unintended ecological and evolutionary consequences for co-circulating pathogens. Future prevention strategies should therefore be evaluated not only according to their direct clinical effectiveness but also according to their long-term impact on antimicrobial resistance across interconnected pathogen populations, and long-term policy decisions should additionally consider both the programmatic efficiency of interventions and their capacity to reshape antimicrobial resistance dynamics through competitive strain replacement.

## Funding

This research is supported by the National Medical Research Council, Singapore, under its HPHSR Clinician Scientist Award (MOH-002048).

## Author contributions

ZW contributed to writing - original draft, software, methodology, formal analysis, and conceptualization. JTL contributed to writing - review & editing, supervision, methodology, formal analysis, and conceptualization. DN and LKW contributed to methodology and conceptualization. All authors contributed to critical review and editing of the manuscript and take responsibility for the decision to submit for publication. JTL is the guarantor and accepts full responsibility for the work and/or the conduct of the study, and controlled the decision to publish. The corresponding author attests that all listed authors meet authorship criteria and that no others meeting the criteria have been omitted.

## Competing interests

The authors declare no competing interests.

## Data and materials availability

All data used to parameterize the model, run the simulations, and generate the findings are included in the Supplementary Information and are publicly available with the model code at the GitHub repository: https://github.com/killingbear999/amr_gonorrhoea. Detailed data sources are included in Methods section (1). Source code to reproduce all our experiments, figures, and analysis is publicly available at the GitHub repository: https://github.com/killingbear999/amr_gonorrhoea.

## Data Availability

All data used to parameterize the model, run the simulations, and generate the findings are included in the Supplementary Information and are publicly available with the model code at the GitHub repository: https://github.com/killingbear999/amr_gonorrhoea.

## Notes

### Competing Interest Statement

The authors have declared no competing interest.

