## Supplementary Information for "The long-term epidemiological impacts of doxycycline post-exposure prophylaxis and vaccination against multidrug-resistant *Neisseria gonorrhoeae*: a mathematical modelling study"

**Section 1: Genetic mechanisms of resistance and mechanisms of antibiotic co-selection**

The vast majority of anorectal and pharyngeal *N. gonorrhoeae* infections are asymptomatic and often self-resolving, and therefore may not come to clinical attention [1]. In addition, many infections remain undiagnosed because regular screening is not universal, even among high-risk populations. Routine surveillance systems capture only diagnosed cases rather than the true underlying incidence. Consequently, gonorrhoea is widely considered to be substantially underdiagnosed and underreported worldwide [2].

In *Neisseria gonorrhoeae*, tetracycline resistance (Tet-R) is predominantly mediated either by the acquisition of plasmid-borne *tetM* determinants, which confer high-level resistance, or via cumulative chromosomal mutations at the *rpsJ*, *mtrR*, and *porB* loci [3]. Conversely, reduced susceptibility or resistance to ceftriaxone (Cef-R) typically arises from the acquisition of mosaic *penA* alleles encoding altered penicillin-binding protein 2 (PBP2), structurally reinforced by compensatory chromosomal mutations within *mtrR* and *porB*. Although the genetic determinants conferring resistance to tetracyclines and extended-spectrum cephalosporins do not share overlapping biochemical pathways, strong empirical genetic associations between these strains have been documented globally [4], [5], [6]. This baseline linkage suggests that widespread exposure to doxycycline - such as through doxycycline post-exposure prophylaxis (doxy-PEP) targeted at syphilis - could substantially expand the existing population-level reservoir of Tet-R *N. gonorrhoeae* strains [7]. This amplified selective pool inherently increases the evolutionary probability for *de novo* Cef-R mutations to emerge within a pre-existing Tet-R genetic background, while simultaneously facilitating the clonal expansion of dual-resistant (Dual-R) strains. Consequently, while doxy-PEP does not exert direct biochemical selective pressure on cephalosporin resistance determinants, it acts as an indirect evolutionary catalyst that can drive up the prevalence of Cef-R *N. gonorrhoeae* through genetic co-selection and ecological niche replacement, particularly where these resistance determinants are structurally linked or co-circulate within identical transmission networks [4].

**Section 2: Strain-specific gonorrhoea transmission model details: Data, methods and calibration**

Source code to reproduce all our experiments, figures, and analysis is publicly available at the GitHub repository: https://github.com/killingbear999/amr_gonorrhoea.

**2.1. Strain-specific gonorrhoea transmission model**

We constructed a strain-specific deterministic, transmission-dynamic compartmental model of gonorrhoea, stratifying the MSM population by infection status and sexual activity level (low vs. high ($j \in L, H$); Fig. 1). The target MSM population is stratified into low-activity and high-activity sexual behaviour classes, with a proportion $q_L$ assigned to the low-activity group and the remainder ($q_H = 1 - q_L$) to the high-activity group. Both strata share identical internal compartmental structures but possess distinct partner-change and transmission transition rates. Demographic renewal is maintained by the continuous entry of individuals into the sexually active population as uninfected, fully susceptible individuals ($U$) at a rate $\alpha$, balanced by a constant background attrition rate across all states due to aging or exit from the active network at a rate $1/\gamma$. Upon exiting, they are removed from the model and no longer contribute to transmission. Because aging is a demographic process independent of infection dynamics and is not an outcome of interest, no separate compartment was included for individuals who leave the population. Clinical progression follows an extended Susceptible-Exposed-Infectious-Treated framework: uninfected ($U$) individuals transition to exposed ($E$) driven by the time-varying strain-specific force of infection $\lambda$ and a corresponding relative fitness $f$, progressing subsequently at a rate $\sigma$ to either asymptomatic ($A$) or symptomatic ($S$) acute infectious states with a probability $1 - \psi$ and $\psi$, respectively. The strain-specific force of infection $\lambda$ is modulated by the corresponding relative fitness parameter $f$, which captures the biological fitness cost associated with antimicrobial resistance. In the absence of antibiotic selection, these fitness costs suppress the transmission advantage of resistant strains, maintaining Cef-R and Dual-R lineages at the low endemic prevalence observed in England surveillance data. Screened individuals including both symptomatic and asymptomatic hosts enter the treated compartment ($T$) prior to recovery at rates

$\mu$ and $\eta$ respectively, while untreated asymptomatic infections can undergo natural clearance ($A \rightarrow U$) at a rate $\nu$. Following successful therapeutic intervention, treated individuals return to the susceptible state ($T \rightarrow U$) at a rate $\rho$ while individuals with Cef-R and Dual-R infections may experience frontline therapeutic treatment failure with a probability of $\phi$ to revert to the actively infectious asymptomatic compartment ($T \rightarrow A$) at a rate $\rho$, establishing a persistent reservoir of transmissible infection. Moreover, the model assumes that natural clearance or successful antibiotic treatment of a gonococcal infection does not confer protective, lasting homotypic immunity, leaving individuals immediately susceptible to reinfection upon recovery [8], [9]. Infections are disaggregated across four co-circulating gonococcal strains characterized by distinct resistance profiles (superscript $k$): tetracycline- and ceftriaxone-susceptible (hereafter referred to as the baseline susceptible) (0), ceftriaxone-resistant ($c$), tetracycline-resistant ($t$), and dual-resistant ($d2$). Ceftriaxone is modelled as it is the sole first-line therapeutic regimen administered in compartment $T$, carrying defined conditional probabilities $\omega_c$ for the emergence of strain-specific resistance during active treatment ($T^0 \rightarrow T^c$ and $T^t \rightarrow T^{d2}$).

The time-varying strain-specific force of infection, defined as the rate at which susceptible individuals with strain $k$ in sexual activity group $j$ acquire infection, $\lambda_j(t)$, is determined by several factors: the annual rate of partner change within each group ($c_j$); the level of assortativity in sexual mixing between groups ($\epsilon$, where $\epsilon$ = 0 represents proportionate mixing and $\epsilon$ = 1 represents fully assortative mixing); the total number of contagious individuals with strain $k$ in group $j$ (${C_j}^k(t)$); the total population in group $j$ ($N_j(t)$); the prevalence of infectious individuals with strain $k$ in each group ($\frac{{C_j}^k(t)}{N_j(t)}$); the proportion of all partnerships in the population that include an individual from group $j$ ($\pi_j(t)$); and the transmission probability. The latter is modelled as a time-varying quantity which increases linearly from the baseline transmission rate ($\beta$) at $t_0$ ($t_0 = 2015$ for England), with an annual increment of $\phi_\beta$. The resulting force of infection is expressed as:

$$ {\lambda_j}^k(t) = c_j\beta\left(1 + \phi_\beta(t - t_0)\right)\left(\epsilon\frac{{C_j}^k(t)}{N_j(t)} + (1-\epsilon)\left(\sum_{i\in\{L,H\}}\pi_i(t)\frac{{C_i}^k(t)}{N_i(t)}\right)\right) \quad (1) $$

where

$$ \pi_j(t) = \frac{c_j N_j(t)}{\sum_{i\in\{L,H\}} c_i\, N_i(t)} \quad (2) $$

$$ {C_j}^k(t) = E_j^k(t) + A_j^k(t) + S_j^k(t) \quad (3) $$

$$ N_j(t) = U_j(t) + \sum_{k\in\{0,c,t,d2\}}\left(E_j^k(t) + A_j^k(t) + S_j^k(t) + T_j^k(t)\right) \quad (4) $$

The screening rate, $\eta_j(t)$, defined as testing in the absence of symptoms and thus applicable to asymptomatic or uninfected individuals and is allowed to vary by sexual activity group. For both groups, the screening rate increases linearly over time, rising by $\phi_\eta$ each year from the initial rate at $t_0$, as follows:

$$ \eta_H(t) = \eta_H(t_0)\left(1 + \phi_\eta(t - t_0)\right) \quad (5) $$

$$ \eta_L(t) = \omega\eta_H(t) \quad (6) $$

To account for differences in behaviour, a factor of $0 < \omega < 1$ is applied, reflecting that individuals in the low-activity group undergo screening less frequently than those in the high-activity group.

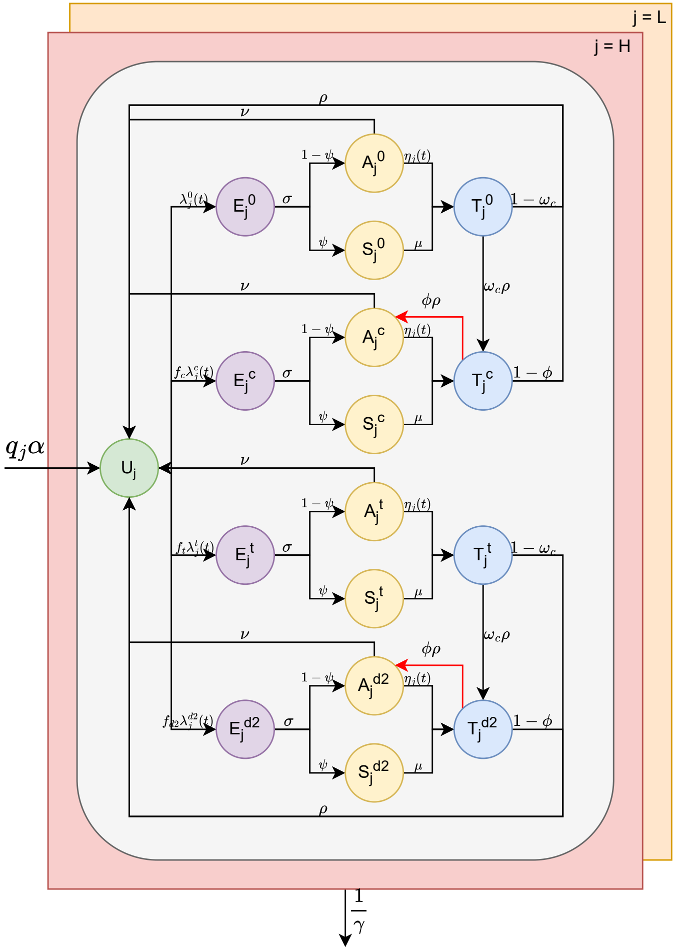


**Fig. 1: Strain-specific gonorrhoea transmission model.** The target MSM population is stratified into low-activity (orange) and high-activity (red) sexual behaviour classes with a proportion $q_L$ assigned to the low-activity group and the remainder ($q_H = 1 - q_L$) to the high-activity group. Both classes share identical internal compartmental structures but possess distinct partner-change and transmission rates; for visual clarity, only inter-compartmental flows for the high-activity cohort (upper layer) are explicitly plotted. Demographic churning is maintained by the continuous entry of individuals into the sexually active population as uninfected, fully susceptible dynamics ($U$) at a rate $\alpha$, balanced by a constant background attrition rate across all states due to aging or exit from the active network at a rate $1/\gamma$. Clinical progression follows an extended

Susceptible-Exposed-Infectious-Treated framework: uninfected ($U$) individuals transition to exposed ($E$) driven by the time-varying strain-specific force of infection $\lambda$ and a corresponding relative fitness $f$, progressing subsequently at a rate $\sigma$ to either asymptomatic ($A$) or symptomatic ($S$) acute infectious states with a probability $1-\psi$ and $\psi$, respectively. Screened individuals including both symptomatic and asymptomatic hosts enter the treated compartment ($T$) prior to recovery at rates $\mu$ and $\eta$ respectively, while untreated asymptomatic infections can undergo natural clearance ($A \rightarrow U$) at a rate $\nu$. Following successful therapeutic intervention, treated individuals return to the susceptible state ($T \rightarrow U$) at a rate $\rho$ while individuals with Cef-R and Dual-R infections experiencing frontline therapeutic treatment failure with a probability of $\phi$ revert to the actively infectious asymptomatic compartment ($T \rightarrow A$; red arrows) at a rate $\rho$, establishing a persistent reservoir of transmissible infection. Infections are disaggregated across four co-circulating gonococcal strains characterized by distinct resistance profiles (superscript $k$): baseline susceptible ($0$), ceftriaxone-resistant ($c$), tetracycline-resistant ($t$), and dual-resistant ($d2$). Ceftriaxone is modelled as the sole first-line therapeutic regimen administered in compartment $T$, carrying defined conditional probabilities $\omega_c$ for the emergence of strain-specific resistance during active treatment ($T^0 \rightarrow T^c$ and $T^t \rightarrow T^{d2}$).

### 2.2. Compartmental model equations

$$\frac{dU_j(t)}{dt} = q_j\alpha + \rho(1-\omega_c)\left(T_j^0(t) + T_j^t(t)\right) + \rho(1-\phi)\left(T_j^c(t) + T_j^{d2}(t)\right)$$
$$+ \nu\left(A_j^0(t) + A_j^c(t) + A_j^t(t) + A_j^{d2}(t)\right) - ({\lambda_j}^0(t) + f_c{\lambda_j}^c(t) + f_t{\lambda_j}^t(t) + f_{d2}{\lambda_j}^{d2}(t) + \frac{1}{\gamma})U_j(t) \quad (7)$$

$$\frac{dE_j^0(t)}{dt} = {\lambda_j}^0(t)U_j(t) - (\sigma + \frac{1}{\gamma})E_j^0(t) \quad (8)$$

$$\frac{dA_j^0(t)}{dt} = \sigma(1-\psi)E_j^0(t) - \left(\nu + \eta_j(t) + \frac{1}{\gamma}\right)A_j^0(t) \quad (9)$$

$$\frac{dS_j^0(t)}{dt} = \sigma\psi E_j^0(t) - (\mu + \frac{1}{\gamma})S_j^0(t) \quad (10)$$

$$\frac{dT_j^0(t)}{dt} = \eta_j(t)A_j^0(t) + \mu S_j^0(t) - \left(\rho + \frac{1}{\gamma}\right)T_j^0(t) \quad (11)$$

$$\frac{dE_j^c(t)}{dt} = f_c{\lambda_j}^c(t)U_j(t) - (\sigma + \frac{1}{\gamma})E_j^c(t) \quad (12)$$

$$\frac{dA_j^c(t)}{dt} = \sigma(1-\psi)E_j^c(t) - \left(\nu + \eta_j(t) + \frac{1}{\gamma}\right)A_j^c(t) + \phi\rho T_j^c(t) \quad (13)$$

$$\frac{dS_j^c(t)}{dt} = \sigma\psi E_j^c(t) - \left(\mu + \frac{1}{\gamma}\right)S_j^c(t) \quad (14)$$

$$\frac{dT_j^c(t)}{dt} = \eta_j(t)A_j^c(t) + \mu S_j^c(t) - \left(\rho + \frac{1}{\gamma}\right)T_j^c(t) + \omega_c\rho T_j^0(t) \quad (15)$$

$$\frac{dE_j^t(t)}{dt} = f_t{\lambda_j}^t(t)U_j(t) - \left(\sigma + \frac{1}{\gamma}\right)E_j^t(t) \quad (16)$$

$$\frac{dA_j^t(t)}{dt} = \sigma(1-\psi)E_j^t(t) - \left(\nu + \eta_j(t) + \frac{1}{\gamma}\right)A_j^t(t) \quad (17)$$

$$\frac{dS_j^t(t)}{dt} = \sigma\psi E_j^t(t) - \left(\mu + \frac{1}{\gamma}\right)S_j^t(t) \quad (18)$$

$$\frac{dT_j^t(t)}{dt} = \eta_j(t)A_j^t(t) + \mu S_j^t(t) - \left(\rho + \frac{1}{\gamma}\right)T_j^t(t) \quad (19)$$

$$\frac{dE_j^{d2}(t)}{dt} = f_{d2}{\lambda_j}^{d2}(t)U_j(t) - (\sigma + \frac{1}{\gamma})E_j^{d2}(t) \quad (20)$$

$$\frac{dA_j^{d2}(t)}{dt} = \sigma(1-\psi)E_j^{d2}(t) - \left(\nu + \eta_j(t) + \frac{1}{\gamma}\right)A_j^{d2}(t) + \phi\rho T_j^{d2}(t) \quad (21)$$

$$\frac{dS_j^{d2}(t)}{dt} = \sigma\psi E_j^{d2}(t) - \left(\mu + \frac{1}{\gamma}\right)S_j^{d2}(t) \quad (22)$$

$$\frac{dT_j^{d2}(t)}{dt} = \eta_j(t)A_j^{d2}(t) + \mu S_j^{d2}(t) - \left(\rho + \frac{1}{\gamma}\right)T_j^{d2}(t) + \omega_c\rho T_j^t(t) \quad (23)$$

### 2.3. Observation processes

First, we compared the observed annual number of gonorrhoea tests and diagnoses (Table 1) in the Genitourinary Medicine Clinic Activity Dataset (GUMCAD) [10] sample, $Z_T(t)$, $Z_D(t)$, with those predicted by our model $Y_T(t)$, $Y_D(t)$ using Negative-Binomial likelihoods to allow for over-dispersion in the observation process relative to a Poisson distribution. If $X \sim$ NegBinom($m$, $\kappa$), with mean $m$ and shape parameter $\kappa$, then:

$$f_X(x|m,\kappa) = \frac{\Gamma(\kappa + x)}{x!\,\Gamma(\kappa)}\left(\frac{\kappa}{\kappa + m}\right)^{\kappa}\left(\frac{m}{\kappa + m}\right)^{x} \quad (24)$$

where the shape parameter characterises the level of clustering or heterogeneity in the observation process, and $\Gamma(\cdot)$ is the Gamma function. Under the Negative-Binomial distribution, the variance of $X$ is $Var[X] = m + m^2/k$.

Specifically, the observation processes of $Z_T(t)$, and $Z_D(t)$ for each strain (i.e., ${Z_D}^0(t)$, ${Z_D}^c(t)$, ${Z_D}^t(t)$, ${Z_D}^{d2}(t)$), are:

$$Z_T(t) \sim NegBinom(Y_T(t), \kappa_T) \tag{25}$$
$$Z_D{}^0(t) \sim NegBinom\left(Y_D{}^0(t), \kappa_T\right) \tag{26}$$
$$Z_D{}^c(t) \sim NegBinom(Y_D{}^c(t), \kappa_T) \tag{27}$$
$$Z_D{}^t(t) \sim NegBinom\left(Y_D{}^t(t), \kappa_T\right) \tag{28}$$
$$Z_D{}^{d2}(t) \sim NegBinom\left(Y_D{}^{d2}(t), \kappa_T\right) \tag{29}$$

where

$$Y_T(t) = \sum_{j\in\{L,H\}} \left( \int_t^{t+1} \begin{matrix} \eta_j(\tau)\left(U_j(\tau) + A_j^0(\tau) + A_j^c(\tau) + A_j^t(\tau) + A_j^{d2}(\tau)\right) \\ + \mu\left(S_j^0(\tau) + S_j^c(\tau) + S_j^t(\tau) + S_j^{d2}(\tau)\right) d\tau \end{matrix} \right) \tag{30}$$

$$Y_D{}^0(t) = \sum_{j\in\{L,H\}} \left( \int_t^{t+1} \rho T_j{}^0(\tau) d\tau \right) \tag{31}$$

$$Y_D{}^c(t) = \sum_{j\in\{L,H\}} \left( \int_t^{t+1} \rho T_j{}^c(\tau) d\tau \right) \tag{32}$$

$$Y_D{}^t(t) = \sum_{j\in\{L,H\}} \left( \int_t^{t+1} \rho T_j{}^t(\tau) d\tau \right) \tag{33}$$

$$Y_D{}^{d2}(t) = \sum_{j\in\{L,H\}} \left( \int_t^{t+1} \rho T_j{}^{d2}(\tau) d\tau \right) \tag{34}$$

with $\tau$ denotes the time variable of integration and $\kappa_T$ denotes the shared shape parameter for GUMCAD data streams.

Second, we compared the observed annual number of symptomatic gonorrhoea diagnoses (Table 1) in the Gonococcal Resistance to Antimicrobials Surveillance Program (GRASP) [11] sample, $Z_S(t)$, with those predicted by our model $Y_S(t)$ using a Beta-Binomial likelihood. If $X$ ~ BetaBinom($n$, $p$, $\kappa$), with size $n$, probability $p$ and over-dispersion $\kappa$, then:

$$f_x(x|n,p,\kappa) = \binom{n}{x} \frac{B(x+a, n-x+b)}{B(a,b)} \tag{35}$$

where $B(\cdot,\cdot)$ is the Beta function and $a = p\left(\frac{1-\kappa}{\kappa}\right)$, $b = (1-p)\left(\frac{1-\kappa}{\kappa}\right)$. Under the Beta-Binomial distribution, the mean and variance of $X$ are $E[X] = np$ and $Var[X] = np(1-p)(1+(n-1)\kappa)$ respectively.

The model-predicted probability that a diagnosis in year $t$ was symptomatic is

$$\frac{Y_S(t)}{Y_S(t) + Y_A(t)} \tag{36}$$

where $Y_S(t)$ and $Y_A(t)$ and the number of symptomatic and asymptomatic diagnoses in year $t$, respectively, as follows:

$$Y_S(t) = \sum_{j\in\{L,H\}} \left( \int_t^{t+1} \mu\left(S_j^0(\tau) + S_j^c(\tau) + S_j^t(\tau) + S_j^{d2}(\tau)\right) d\tau \right) \tag{37}$$

$$Y_A(t) = \sum_{j\in\{L,H\}} \left( \int_t^{t+1} \eta_j(\tau)\left(A_j^0(\tau) + A_j^c(\tau) + A_j^t(\tau) + A_j^{d2}(\tau)\right) d\tau \right) \tag{38}$$

Specifically, we modelled the number of symptomatic diagnoses in GRASP, accounting for the size of the GRASP sample, $Z_R(t)$, and fitted the shape parameter for the GRASP dataset, $\kappa_S$, within the Bayesian framework:

$$Z_S(t) \sim BetaBinom\left(Z_R(t), \frac{Y_S(t)}{Y_S(t) + Y_A(t)}, \kappa_S\right) \tag{39}$$

The overall likelihood of the data given the modelled trajectories produced by parameter set Θ was calculated as the product of the likelihoods of the six data streams in each year $t$ = 2011 ($t_0$), …, 2019, 2022, 2023, 2024 ($t_{max}$) for England ($t = 2020, 2021$ were excluded for England for all six data streams), as follows:

$$L(Z|\Theta) = \prod_{t=t_0}^{t_{max}} f_{Z_T}(Z_T(t)|\Theta) f_{Z_D{}^c}(Z_D{}^c(t)|\Theta) f_{Z_D{}^{d2}}\left(Z_D{}^{d2}(t)\middle|\Theta\right) \prod_{t=2015}^{2022} f_{Z_D{}^0}\left(Z_D{}^0(t)\middle|\Theta\right) f_{Z_D{}^t}\left(Z_D{}^t(t)\middle|\Theta\right) \prod_{t=2013}^{t_{max}} f_{Z_S}(Z_S(t)|Z_R(t), \Theta) \tag{40}$$

Gonorrhoea can infect multiple anatomical sites (i.e., the rectum, pharynx, and urethra) each associated with distinct probabilities of symptom development and transmission potential [12]. Because available surveillance data are not disaggregated by infection site, model parameters represent site-averaged values. The model further assumes that infection does not confer lasting natural immunity [8], [9].

**Table 1. Annual gonorrhoea diagnoses, tests, symptomatic diagnoses, asymptomatic diagnoses, percentage ceftriaxone-resistant, and percentage tetracycline-resistant among MSM in England.** Doxycycline susceptibility cannot be directly assessed because doxycycline MICs were not routinely available. Therefore, we used tetracycline MIC > 1 mg/L among MSM isolates as the primary proxy for doxycycline non-susceptibility, consistent with surveillance practice. High-level plasmid-mediated resistance (tetM), typically associated with tetracycline MIC ≥ 8 mg/L, was available only for the general population; these data were used solely for sensitivity analyses to bound the prevalence of high-level doxycycline resistance in MSM.

| Year | Number of diagnoses | Number of tests | Number of MSM in GRASP report | Number of symptomatic diagnoses in GRASP report | Number of asymptomatic diagnoses in GRASP report | Percentage ceftriaxone-resistant in MSM (MIC > 0.125mg/L) | Percentage tetracycline-resistant in MSM (MIC > 1mg/L) | Percentage tetracycline-resistant (MIC ≥ 8mg/L) |
|---|---|---|---|---|---|---|---|---|
| 2011 | 7797 | 86472 | - | - | - | 0.0 | - | - |
| 2012 | 10690 | 99558 | - | - | - | 0.0 | - | 9.5 |
| 2013 | 13579 | 114282 | 1090 | 402 | 191 | 0.0 | 92.5[1] | 11.2 |
| 2014 | 18017 | 146991 | 1073 | 573 | 361 | 0.0 | 91.5[*] | - |
| 2015 | 22042 | 162071 | 1226 | 454 | 358 | 0.0 | 41.6 | - |
| 2016 | 17297 | 170936 | 821 | 366 | 325 | 0.0 | 45.1 | - |
| 2017 | 21209 | 190941 | 839 | 355 | 327 | 0.0 | 54.4 | 43.7 |
| 2018 | 26748 | 228103 | 799 | 361 | 328 | 0.0 | 63.6 | 43.2 |
| 2019 | 33634 | 268605 | 971 | 408 | 419 | 0.0 | 75.0 | 45.2 |
| 2020[**] | 24936 | 219721 | 785 | 410 | 265 | 0.0 | 79.2 | 30.7 |
| 2021[**] | 24830 | 260778 | 894 | 411 | 483 | 0.0 | 86.0 | 33.6 |
| 2022 | 36933 | 323012 | 852 | 367 | 481 | 0.0 | 68.5 | 18.3 |
| 2023 | 40344 | 364015 | 1052 | 406 | 646 | 0.0 | 85.9[***] | 16.8 |
| 2024 | 38647 | 372371 | 971 | 437 | 530 | 0.0 | - | 30.1 |

[1] MIC ≥ 2mg/L
[**] Data from 2020 - 2021 were excluded due to sampling biases during COVID-19.
[***] MIC > 0.5mg/L

### 2.4. Calibration process

The strain-specific gonorrhoea transmission model (Fig. 1) was calibrated using annual gonorrhoea diagnoses, tests, symptomatic diagnoses, asymptomatic diagnoses, percentage ceftriaxone-resistant, and percentage tetracycline-resistant data among MSM (Table 1) and sexual behaviour data (Table 2) from England within a Bayesian framework.

**Table 2: Fixed model parameters (England): notation, definitions, source of estimates.**

| | Definition | Value | Source |
|---|---|---|---|
| $N(t_0)$ | Initial population size of England MSM | 600,000 | [13] |
| $\alpha$ | Annual population entrance (at age 15) | 12,000 | [13] |
| $\gamma$ | Years spent in the sexually-active population | 50 | Ages 15-65 |
| $q_L$ | Proportion of the population in group $L$ | 0.85 | [13] |
| $q_H$ | Proportion of the population in group $H$ | 0.15 | $1 - q_L$ |
| $c_L$ | Annual rate of partner change in group $L$ | 0.6 | [13] |
| $c_H$ | Annual rate of partner change in group $H$ | 15.6 | [13] |

England serves as an ideal epidemiological setting for evaluating the evolutionary dynamics of *N. gonorrhoeae* AMR under dual intervention pressures. The jurisdiction maintains world-class, comprehensive national surveillance registries, specifically Genitourinary Medicine Clinic Activity Dataset (GUMCAD) [10] and Gonococcal Resistance to Antimicrobials Surveillance Programme (GRASP) [11], which provide high-resolution, longitudinal data on sexually transmitted infection (STI) testing volumes, clinical presentations, and strain-specific resistance profiles. Furthermore, transmission in the English MSM population is characterized by a relatively high but not yet saturated baseline prevalence of tetracycline resistance [14] and a complete absence of sustained and domestically acquired ceftriaxone-resistant *N. gonorrhoeae* clusters to date [15], but there are rising pressures on frontline ceftriaxone susceptibility reported in England since 2021 [16]. This context has the epidemiological data required for model calibration and evaluating targeted public health interventions.

For epidemiological data (Table 1), data from 2020 and 2021 were excluded from both model calibration and simulation due to substantial disruptions in contact rates associated with the COVID-19 pandemic. During this period, sexual behaviour, healthcare access, testing practices, clinic attendance, and reporting systems were markedly altered by lockdown measures, service reallocation, and changes in care-seeking behaviour. These disruptions resulted in surveillance data that do not reflect the underlying natural transmission dynamics assumed in our model framework. Because the transmission model does not explicitly incorporate COVID-19-related behavioural restrictions or service interruptions, including these years in either the likelihood (calibration) or prediction phase would potentially influence model calibration. This could bias parameter estimates, distort inferred transmission trends, and reduce predictive validity for post-pandemic scenarios. Excluding 2020-2021 therefore ensures internal consistency between model structure and observed data, yielding parameter estimates that better represent stable transmission conditions and providing more reliable projections for future, non-pandemic epidemiological settings.

We specified priors for the model parameters to reflect existing knowledge and uncertainty about their plausible ranges, as detailed in Table 3. For parameters with high uncertainty, whose values are best informed by the data, we used wide Uniform priors. These include: the probability of transmission per partnership ($\beta$); the annual increase in transmission risk behaviour ($\phi_\beta$); the level of assortativity in sexual mixing ($\epsilon$); the initial rate of asymptomatic screening in group $H$ ($\eta_H(t_0)$); the annual increase in asymptomatic screening rate ($\phi_\eta$); the probability that incident infection is symptomatic ($\psi$); and the shape parameter of communicable disease surveillance data ($\kappa_T$ and $\kappa_S$). The remaining parameters, related to the natural history of gonorrhoea, were assigned LogNormal priors. For all fitted parameters, we imposed reflective boundaries to ensure that proposed values remained both mathematically valid and epidemiologically plausible. Specifically, we employed Hamiltonian Monte Carlo (HMC) via the RStan package (version 2.32.7) in R (version 4.5.0), running six chains of 2,000 iterations each, with the first 1,000 discarded as burn-in. The notation, definitions, prior distributions and posterior estimates of the fitted model parameters are presented in Table 3.

Fig. 2 presents the observed and simulated *N. gonorrhoeae* epidemic trajectories among MSM in England, disaggregated by resistance strain. The aggregate volume of diagnosed incident infections exhibits a progressive increase across the historical baseline period. When disaggregated by specific resistance strains, baseline susceptible and Tet-R strains drive this overall escalation, both exhibiting a progressive linear increase. Conversely, the transmission volumes for Cef-R and Dual-R strains remain stably suppressed at near-zero endemic levels throughout the baseline timeline.

**Table 3: Fitted parameters: notation, definitions, prior distributions, and posterior estimates.** Transition rate parameters ($\theta \in \{\sigma, \mu, \rho, \nu\}$) are presented in an annual basis, giving a mean time to transition of $365/\theta$ days. Posterior estimates are taken as the median, with 95% credible interval presented in parentheses.

| | Definition | Prior distribution | Parameter bound | Posterior estimate | Source[2] |
|---|---|---|---|---|---|
| $\beta$ | Probability of transmission per-partnership | U[0, 1] | [0, 1] | 0.356 (0.199 – 0.818) | [17] |
| $\phi_\beta$ | Annual increase in transmission risk behaviour | U[0, 1] | [0, 0.5] | 0.0655 (0.00336 – 0.430) | [17] |
| $\epsilon$ | Level of assotativity in sexual mixing | U[0, 1] | [0, 1] | 0.576 (0.0262 – 0.982) | [17] |
| $\sigma$ | Rate of leaving incubation period | logN[4.60, 0.29] | [0, 1000] | 99.185 (55.912 – 171.901) | [17] |
| $\psi$ | Probability that incident infection is symptomatic | U[0, 1] | [0, 1] | 0.117 (0.0299 – 0.399) | [17] |
| $\mu$ | Rate of seeking treatment due to symptom | logN[5.38, 0.44] | [0, 2000] | 215.691 (90.767 – 516.474) | [17] |
| $\eta_H(t_0)$ | Initial rate of asymptomatic screening in group $H$ | U[0, 4] | [0, 2] | 0.401 (0.131 – 1.381) | [17] |
| $\omega$ | Ratio of screening rate in group $L$ vs $H$ | logN[-0.87, 0.39] | [0.1, 1] | 0.424 (0.196 – 0.831) | [17] |
| $\phi_\eta$ | Annual increase in screening rate | U[0, 1] | [0, 0.3] | 0.115 (0.00464 – 0.289) | [17] |
| $\rho$ | Rate of recovery after treatment | logN[3.99, 0.11] | [0, 200] | 54.083 (43.511 – 67.040) | [17] |
| $\nu$ | Rate of natural recovery | logN[1.13, 0.34] | [0, 50] | 2.919 (1.490 – 5.535) | [17] |
| $\phi$ | Treatment failure rate for ceftriaxone- or dual resistant infections treated with ceftriaxone | Γ[0.001, 1000] | [0, 0.2] | 0.0000232 (0.00000119 – 0.00124) | [18] |
| $f_c$ | Relative fitness of ceftriaxone-resistant bacteria, compared with baseline susceptible | logN[-0.0204, 0.02] | [0.7, 1.1] | 0.980 (0.942 – 1.018) | [19] |
| $f_t$ | Relative fitness of tetracycline-resistant bacteria, compared with baseline susceptible | logN[-0.0204, 0.02] | [0.7, 1.1] | 0.990 (0.955 – 1.03) | [19] |
| $f_{d2}$ | Relative fitness of dual resistant bacteria, compared with baseline susceptible | logN[-0.0408, 0.02] | [0.5, 1] | 0.959 (0.922 – 0.992) | [19] |
| $w_c$ | Probability of emergence of ceftriaxone resistance upon treatment with ceftriaxone | logN[$\log 10^{-8}$, 0.01] | [$10^{-10}$, $10^{-5}$] | 0.0000000100 (0.0000000098 – 0.0000000102) | [19] |
| $w_t$ | Probability of emergence of tetracycline resistance upon taking doxy-PEP for syphilis after gonorrhoea infection | Γ[0.001, 1000] | [$10^{-6}$, $10^{-2}$] | 0.0000268 (0.00000120, 0.00112) | [19] |
| $\kappa_T$ | Shape parameter of GUMCAD dataset | U[0,1] | [0.05, 1] | 0.944 (0.741 – 0.998) | - |
| $\kappa_S$ | Shape parameter of GRASP dataset | U[0,1] | [0.05, 1] | 0.876 (0.508 – 0.995) | - |

[2] For prior distributions and parameter bounds.

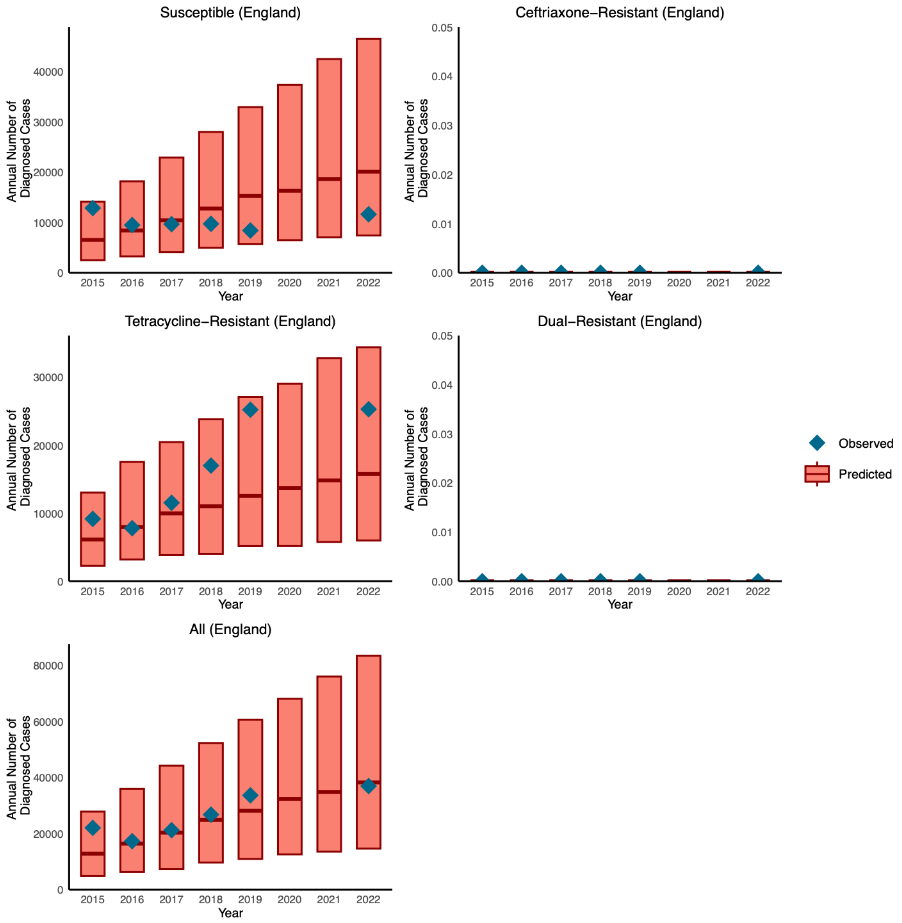


**Fig. 2: Observed and simulated *N. gonorrhoeae* epidemic trajectories among MSM in England, disaggregated by resistance strain (baseline susceptible, Tet-R, Cef-R, Dual-R, and total diagnosed cases, respectively).** Each box represents the posterior distribution, with the median indicated by a horizontal line and the interquartile range represented by the upper and lower edges of the box. Observed annual gonorrhoea incidence data are shown as diamonds.

#### 2.5. Calibration convergence statistics

Convergence diagnostics - including traceplots (see Fig. 3), marginal posterior distributions, effective sample size (ESS), and the Gelman-Rubin (GR) statistic (see Table 4) - were used to ensure robust parameter estimation.

**Table 4. The ESS and GR diagnostic of posterior parameter estimates.**

| | England | |
|---|---|---|
| Parameter | ESS | GR |
| $\beta$ | 1,673 | 1.00 |
| $\phi_\beta$ | 2,895 | 1.00 |
| $\epsilon$ | 6,033 | 1.00 |
| $\sigma$ | 7,798 | 1.00 |
| $\psi$ | 2,079 | 1.00 |
| $\mu$ | 5,871 | 1.00 |
| $\eta_H(t_0)$ | 1,849 | 1.00 |
| $\omega$ | 3,855 | 1.00 |
| $\phi_\eta$ | 5,772 | 1.00 |
| $\rho$ | 6,716 | 1.00 |
| $\nu$ | 4,922 | 1.00 |
| $\phi$ | 5,333 | 1.00 |
| $f_c$ | 7,312 | 1.00 |
| $f_t$ | 5,385 | 1.00 |
| $f_{d2}$ | 4,865 | 1.00 |
| $w_c$ | 8,899 | 1.00 |
| $w_t$ | 5,644 | 1.00 |
| $\kappa_T$ | 7,986 | 1.00 |
| $\kappa_S$ | 9,970 | 1.00 |

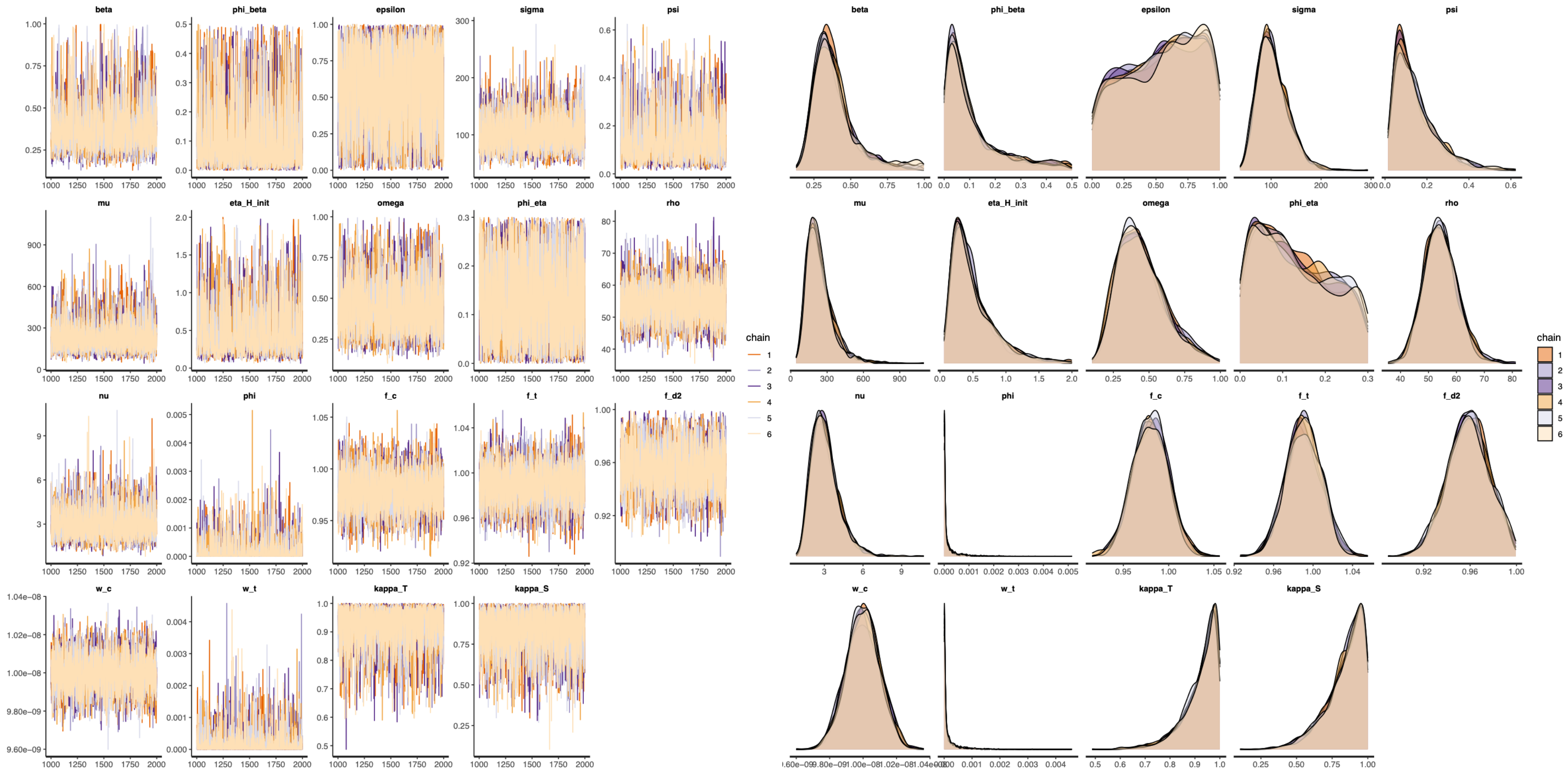


**Fig. 3: Traceplots (left column) and marginal posterior densities (right column) of parameter estimates.** Traceplots show MCMC samples from five chains, each with 2,000 iterations, with the first 1,000 discarded as burn-in. Coloured lines represent individual chains.

**Section 3: Dual-intervention model for gonorrhoea: Methods and simulation**

**3.1. Model of dual intervention**

We extended the fitted transmission-dynamic model (Fig. 1) to include doxy-PEP and vaccination, allowing for different levels of behavioural patterns such as uptake rate and adherence rate (see Table 5). For the dual-intervention model, see Fig. 4, demographic churning is maintained by the continuous entry of individuals into the sexually active population as uninfected, fully susceptible individuals ($U$) within the non-intervention stratum ($N$) at a rate $\alpha$, balanced by a constant background attrition rate across all states due to aging or exit from the active network at a rate $1/\gamma$. The population is split into four distinct sub-populations based on coverage and uptake status: non-intervention ($N$), doxy-PEP standalone intervention ($D$), vaccination standalone intervention ($V$), and combined dual-interventions ($M$). With the four strata, the time-varying strain-specific force of infection is then defined similarly to Equation (1), but with different total number of contagious individuals in group $j$ ($C_j(t)$) and total population in group $j$ ($N_j(t)$), as follows:

$$\lambda_j{}^k(t) = c_j\beta\left(1 + \phi_\beta(t - t_0)\right)\left(\epsilon\frac{C_j{}^k(t)}{N_j(t)} + (1 - \epsilon)\left(\sum_{i\in\{L,H\}} \pi_i(t)\frac{C_i{}^k(t)}{N_i(t)}\right)\right) \tag{41}$$

where

$$\pi_j(t) = \frac{c_j N_j(t)}{\sum_{i\in\{L,H\}} c_i\, N_i(t)} \tag{42}$$

$$C_j{}^k(t) = \sum_{i\in\{N,D,V,M\}} \left(E_{i,j}^k(t) + A_{i,j}^k(t) + S_{i,j}^k(t)\right) \tag{43}$$

$$N_j(t) = \sum_{i\in\{N,D,V,M\}} \left(U_{i,j}(t) + \sum_{k\in\{0,c,t,d2\}} \left(E_{i,j}^k(t) + A_{i,j}^k(t) + S_{i,j}^k(t) + T_{i,j}^k(t)\right)\right) \tag{44}$$

Within each stratum ($i \in \{N, D, V, M\}$), clinical progression follows an extended Susceptible-Exposed-Infectious-Treated framework: uninfected ($U$) individuals transition to exposed ($E$) driven by the time-varying strain-specific force of infection $\lambda$ and a corresponding relative fitness $f$, progressing subsequently to either asymptomatic ($A$) with a probability $1 - \psi$ or symptomatic ($S$) with a probability $\psi$ acute infectious states at a rate $\sigma$. Screened individuals (including both symptomatic at a rate $\mu$ and asymptomatic hosts at a screening-dependent rate $\eta$) enter the treated compartment ($T$) prior to recovery, while untreated asymptomatic infections can undergo natural clearance ($A \to U$) at a rate $\nu$. Following successful therapeutic intervention, treated individuals return to the susceptible state ($T \to U$) at a rate $\rho$, while individuals with Cef-R and Dual-R infections may experience frontline therapeutic treatment failure with a probability of $\phi$ to revert to the actively infectious asymptomatic compartment ($T \to A$; red arrows) at a rate $\rho$, establishing a persistent reservoir of transmissible infection. For analytical simplicity, intervention initiation is restricted to the uninfected ($U$) state.

Infections within each sub-population are disaggregated across four co-circulating gonococcal strains characterized by distinct resistance profiles (superscript $k$): baseline susceptible ($0$), ceftriaxone-resistant ($c$), tetracycline-resistant ($t$), and dual-resistant ($d2$). Ceftriaxone is modelled as the sole first-line therapeutic regimen administered in compartment $T$, carrying defined conditional probabilities $\omega_c$ for the emergence of strain-specific resistance during active treatment ($T^0 \to T^c$ and $T^t \to T^{d2}$). The directional selective and protective forces of the interventions modulate the effective force of infection ($\lambda$) via state-specific modifiers: Doxy-PEP and vaccination together reduce the effective force of infection ($U \to E^0$ and $U \to E^c$; yellow arrows in stratum $V$) by $1 - e_{vd}$. Vaccination standalone intervention exerts an evolutionary neutral, transmission-blocking effect, reducing the effective force of infection uniformly across all four bacterial strains ($U \to E^0$, $U \to E^c$, $U \to E^t$, and $U \to E^{d2}$; blue arrows in strata $V$ and $M$) by $e_v$. Doxy-PEP standalone intervention suppresses incoming baseline susceptible and ceftriaxone-resistant exposures ($U \rightarrow E^0$ and $U \rightarrow E^c$; green arrows in stratum $D$) by $e_d$, while leaving the transmissibility of tetracycline-resistant and dual-resistant lineages unaltered ($U \rightarrow E^t$ and $U \rightarrow E^{d2}$; black arrows in strata $N$ and $D$). Crucially, active doxy-PEP exposure introduces a non-zero probability for the *de novo* selection of tetracycline resistance during prophylaxis (dotted red arrows in strata $D$ and $V$) with a probability $\omega_t$. Transitions between intervention states are governed by: (**1**) Doxy-PEP initiation drives flows from $N \rightarrow D$ and $M \rightarrow V$ (pink arrows) with a probability $p_d$, (**2**) vaccine initiation shifts individuals from $N \rightarrow M$ and $D \rightarrow V$ (dark green arrows) with a probability $p_v$. (**3**) Conversely, the cessation or discontinuation of doxy-PEP reverts individuals from $D \rightarrow N$ and $V \rightarrow M$ (dark blue arrows) at a rate $\xi_n$. (**4**) Temporal waning of vaccine-induced protection returns individuals from $V \rightarrow D$ and $M \rightarrow N$ (dark orange arrows) at a rate $\xi_d$.

A key structural boundary of our model is the assumption that resistance to tetracycline and resistance to ceftriaxone are governed by independent genetic loci. Consequently, we assume no direct genetic linkage or intrinsic biological co-selection mechanisms between these resistance determinants; any observed epidemiological co-selection is driven exclusively by ecological replacement and population-level antibiotic selection pressures. This modelling choice reflects a pragmatic abstraction necessitated by current surveillance data limitations. While high-resolution genomic data tracking the precise intra-strain plasmidic or chromosomal linkage of these specific determinants were not systematically available across the full calibration timeline, empirical evidence suggests that strain-specific multi-drug resistance in *N. gonorrhoeae* predominantly emerges through sequential, unlinked structural mutations under distinct clinical selection pressures [20].

**Table 5: Model parameters used in scenario analysis.**

| | Definition | Value | Source |
|---|---|---|---|
| $e_d$ | Doxy-PEP efficacy against gonorrhoea infection | 55% | [21], [22] |
| $e_v$ | Vaccination efficacy against gonorrhoea infection | 40% | [23] |
| $e_{vd}$ | Combined efficacy of doxy-PEP and vaccination against gonorrhoea infection | 73%[3] | - |
| $p_d$ | Uptake of doxy-PEP for syphilis | 0-100% | - |
| $p_v$ | Uptake of vaccination for gonorrhoea | 0-100% | - |
| $\xi_n$ | Discontinuation rate of doxy-PEP for syphilis | 0.362 | [24] |
| $\xi_d$ | Waning rate of vaccination | 0.20[4] | [25], [26] |

[3] Assumed each intervention reduces infection risk independently such that their combined efficacy is modelled multiplicatively, i.e., $e_{vd} = 1 - (1 - e_d)(1 - e_v)$.
[4] A booster dose may need to be further evaluated and considered in adolescents, given evidence of waning protection against gonorrhoea beginning around 36 months after vaccination [25]. Moderate protection against both first and subsequent gonococcal infections has been observed for up to 5 years following vaccination, although protection appears to decline over time, with waning becoming evident after 5 years [26].

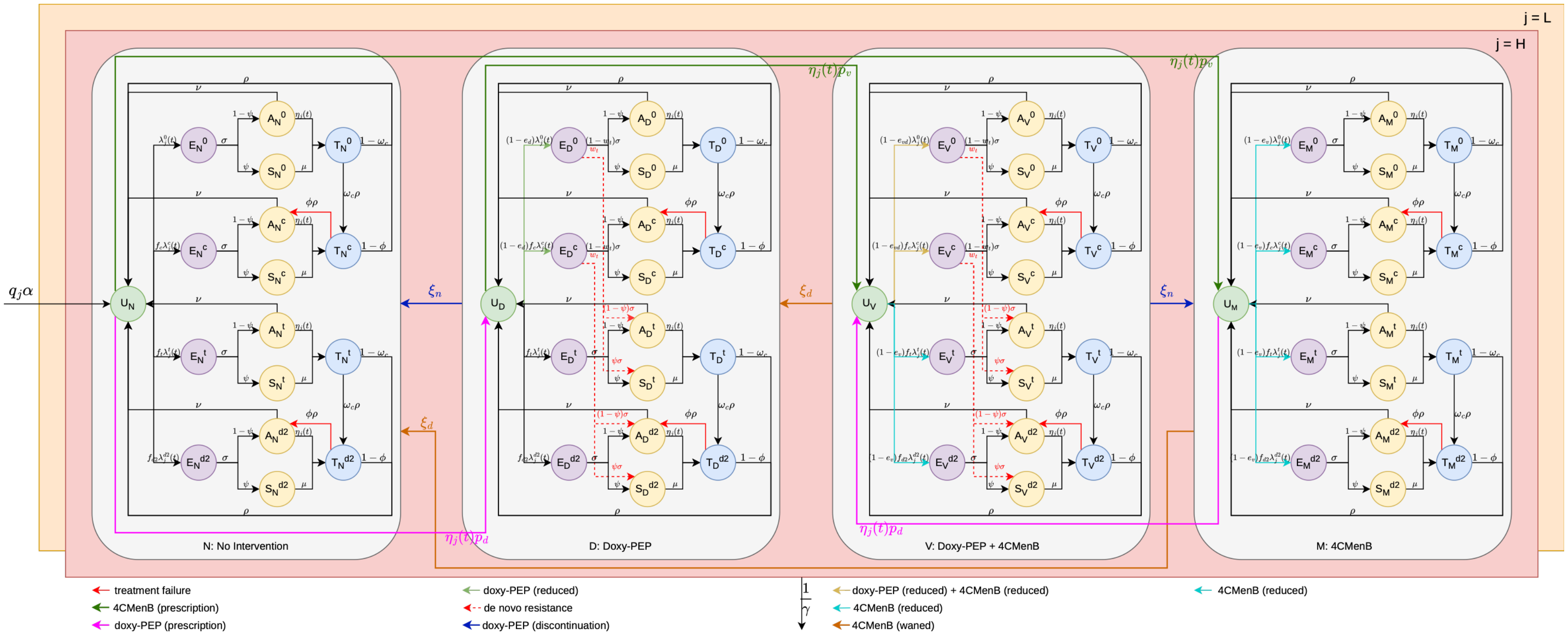


**Fig. 4: Compartmental structure and transmission architecture of the strain-stratified *N. gonorrhoeae* model under dual intervention pressures.** The schematic illustrates the transmission dynamics and evolutionary pathways of *N. gonorrhoeae* under targeted doxy-PEP and vaccination. The target MSM population is stratified into low-activity (orange) and high-activity (red) sexual behaviour classes. Both classes share identical internal compartmental structures but possess distinct partner-change and transmission transition rates; for visual clarity, only inter-compartmental flows for the high-activity cohort (upper layer) are explicitly plotted. Demographic churning is maintained by the continuous entry of individuals into the sexually active population as uninfected, fully susceptible dynamics ($U$) within the non-intervention stratum ($N$) at a rate $\alpha$, balanced by a constant background attrition rate across all states due to aging or exit from the active network at a rate $1/\gamma$. The population is split into four distinct sub-populations based on coverage and uptake status: non-intervention ($N$), doxy-PEP standalone intervention ($D$), vaccination standalone intervention ($V$), and combined dual-interventions ($M$). Within each stratum ($i \in \{N, D, V, M\}$), clinical progression follows an extended Susceptible-Exposed-Infectious-Treated framework: uninfected ($U$) individuals transition to exposed ($E$) driven by the time-varying strain-specific force of infection $\lambda$ and a corresponding relative fitness $f$, progressing subsequently at a rate $\sigma$ to either asymptomatic ($A$) or symptomatic ($S$) acute infectious states with a probability $1-\psi$ and $\psi$, respectively. Screened individuals including both symptomatic and

asymptomatic hosts enter the treated compartment ($T$) prior to recovery at rates $\mu$ and $\eta$ respectively, while untreated asymptomatic infections can undergo natural clearance ($A \rightarrow U$) at a rate $\nu$. Following successful therapeutic intervention, treated individuals return to the susceptible state ($T \rightarrow U$) at a rate $\rho$ while individuals with Cef-R and Dual-R infections experiencing frontline therapeutic treatment failure with a probability of $\phi$ revert to the actively infectious asymptomatic compartment ($T \rightarrow A$; red arrows) at a rate $\rho$, establishing a persistent reservoir of transmissible infection. For analytical simplicity, intervention initiation is restricted to the uninfected ($U$) state. Infections within each sub-population are disaggregated across four co-circulating gonococcal strains characterized by distinct resistance profiles (superscript $k$): baseline susceptible ($0$), ceftriaxone-resistant ($c$), tetracycline-resistant ($t$), and dual-resistant ($d2$). Ceftriaxone is modelled as the sole first-line therapeutic regimen administered in compartment $T$, carrying defined conditional probabilities $\omega_c$ for the emergence of strain-specific resistance during active treatment ($T^0 \rightarrow T^c$ and $T^t \rightarrow T^{d2}$). The directional selective and protective forces of the interventions modulate the effective force of infection ($\lambda$) via state-specific modifiers: Doxy-PEP and vaccination together reduce the effective force of infection ($U \rightarrow E^0$ and $U \rightarrow E^c$; yellow arrows in stratum $V$) by $1 - e_{vd}$. Vaccination standalone intervention exerts an evolutionary neutral, transmission-blocking effect, reducing the effective force of infection uniformly across all four bacterial strains ($U \rightarrow E^0$, $U \rightarrow E^c$, $U \rightarrow E^t$, and $U \rightarrow E^{d2}$; blue arrows in strata $V$ and $M$) by $e_v$. Doxy-PEP standalone intervention acts as an ecological filter, suppressing incoming baseline susceptible and ceftriaxone-resistant exposures ($U \rightarrow E^0$ and $U \rightarrow E^c$; green arrows in stratum $D$) by $e_d$, while leaving the transmission velocity of tetracycline-resistant and dual-resistant lineages unaltered ($U \rightarrow E^t$ and $U \rightarrow E^{d2}$; black arrows in strata $N$ and $D$). Crucially, active doxy-PEP exposure introduces a non-zero probability for the *de novo* selection of tetracycline resistance during prophylaxis (dotted red arrows in strata $D$ and $V$) with a probability $\omega_t$. Policy transitions and immunity dynamics dictate movement between the four coverage layers: doxy-PEP initiation drives flows from $N \rightarrow D$ and $M \rightarrow V$ (pink arrows) with a probability $p_d$, while vaccine rollout shifts individuals from $N \rightarrow M$ and $D \rightarrow V$ (dark green arrows) with a probability $p_v$. Conversely, the cessation or discontinuation of doxy-PEP reverts individuals from $D \rightarrow N$ and $V \rightarrow M$ (dark blue arrows) at a rate $\xi_n$, whereas the temporal waning of vaccine-induced protection returns individuals from $V \rightarrow D$ and $M \rightarrow N$ (dark orange arrows) at a rate $\xi_d$.

#### 3.2. Compartmental model equations

For compartmental model equations, subscript denotes stratum $i \in \{N, D, V, M\}$ and risk group $j \in \{H, L\}$, separated by comma, superscript denotes strain $k \in \{0, c, t, d2\}$.

a. For no-intervention (N):

$$\begin{aligned}\frac{dU_{N,j}(t)}{dt} &= q_j\alpha + \rho(1-\omega_c)\left(T^0_{N,j}(t)+T^t_{N,j}(t)\right) + \rho(1-\phi)\left(T^c_{N,j}(t)+T^{d2}_{N,j}(t)\right) \\ &+ \nu\left(A^0_{N,j}(t)+A^c_{N,j}(t)+A^t_{N,j}(t)+A^{d2}_{N,j}(t)\right) - (\lambda_j{}^0(t) + f_c\lambda_j{}^c(t) + f_t\lambda_j{}^t(t) + f_{d2}\lambda_j{}^{d2}(t) \\ &+ (p_d+p_v)\eta_j(t) + \frac{1}{\gamma})U_{N,j}(t) + \xi_n U_{D,j}(t) + \xi_d U_{M,j}(t)\end{aligned} \tag{45}$$

$$\frac{dE^0_{N,j}(t)}{dt} = \lambda_j{}^0(t)U_{N,j}(t) - (\sigma + \frac{1}{\gamma})E^0_{N,j}(t) + \xi_n E^0_{D,j}(t) + \xi_d E^0_{M,j}(t) \tag{46}$$

$$\frac{dA^0_{N,j}(t)}{dt} = \sigma(1-\psi)E^0_{N,j}(t) - \left(\nu + \eta_j(t) + \frac{1}{\gamma}\right)A^0_{N,j}(t) + \xi_n A^0_{D,j}(t) + \xi_d A^0_{M,j}(t) \tag{47}$$

$$\frac{dS^0_{N,j}(t)}{dt} = \sigma\psi E^0_{N,j}(t) - (\mu + \frac{1}{\gamma})S^0_{N,j}(t) + \xi_n S^0_{D,j}(t) + \xi_d S^0_{M,j}(t) \tag{48}$$

$$\frac{dT^0_{N,j}(t)}{dt} = \eta_j(t)A^0_{N,j}(t) + \mu S^0_{N,j}(t) - \left(\rho + \frac{1}{\gamma}\right)T^0_{N,j}(t) + \xi_n T^0_{D,j}(t) + \xi_d T^0_{M,j}(t) \tag{49}$$

$$\frac{dE^c_{N,j}(t)}{dt} = f_c\lambda_j{}^c(t)U_{N,j}(t) - (\sigma + \frac{1}{\gamma})E^c_{N,j}(t) + \xi_n E^c_{D,j}(t) + \xi_d E^c_{M,j}(t) \tag{50}$$

$$\frac{dA^c_{N,j}(t)}{dt} = \sigma(1-\psi)E^c_{N,j}(t) - \left(\nu + \eta_j(t) + \frac{1}{\gamma}\right)A^c_{N,j}(t) + \xi_n A^c_{D,j}(t) + \xi_d A^c_{M,j}(t) + \phi\rho T^c_{N,j}(t) \tag{51}$$

$$\frac{dS^c_{N,j}(t)}{dt} = \sigma\psi E^c_{N,j}(t) - \left(\mu + \frac{1}{\gamma}\right)S^c_{N,j}(t) + \xi_n S^c_{D,j}(t) + \xi_d S^c_{M,j}(t) \tag{52}$$

$$\frac{dT^c_{N,j}(t)}{dt} = \eta_j(t)A^c_{N,j}(t) + \mu S^c_{N,j}(t) - \left(\rho + \frac{1}{\gamma}\right)T^c_{N,j}(t) + \xi_n T^c_{D,j}(t) + \xi_d T^c_{M,j}(t) + \omega_c\rho T^0_{N,j}(t) \tag{53}$$

$$\frac{dE^t_{N,j}(t)}{dt} = f_t\lambda_j{}^t(t)U_{N,j}(t) - \left(\sigma + \frac{1}{\gamma}\right)E^t_{N,j}(t) + \xi_n E^t_{D,j}(t) + \xi_d E^t_{M,j}(t) \tag{54}$$

$$\frac{dA^t_{N,j}(t)}{dt} = \sigma(1-\psi)E^t_{N,j}(t) - \left(\nu + \eta_j(t) + \frac{1}{\gamma}\right)A^t_{N,j}(t) + \xi_n A^t_{D,j}(t) + \xi_d A^t_{M,j}(t) \tag{55}$$

$$\frac{dS^t_{N,j}(t)}{dt} = \sigma\psi E^t_{N,j}(t) - \left(\mu + \frac{1}{\gamma}\right)S^t_{N,j}(t) + \xi_n S^t_{D,j}(t) + \xi_d S^t_{M,j}(t) \tag{56}$$

$$\frac{dT^t_{N,j}(t)}{dt} = \eta_j(t)A^t_{N,j}(t) + \mu S^t_{N,j}(t) - \left(\rho + \frac{1}{\gamma}\right)T^t_{N,j}(t) + \xi_n T^t_{D,j}(t) + \xi_d T^t_{M,j}(t) \tag{57}$$

$$\frac{dE^{d2}_{N,j}(t)}{dt} = f_{d2}\lambda_j{}^{d2}(t)U_{N,j}(t) - (\sigma + \frac{1}{\gamma})E^{d2}_{N,j}(t) + \xi_n E^{d2}_{D,j}(t) + \xi_d E^{d2}_{M,j}(t) \tag{58}$$

$$\frac{dA^{d2}_{N,j}(t)}{dt} = \sigma(1-\psi)E^{d2}_{N,j}(t) - \left(\nu + \eta_j(t) + \frac{1}{\gamma}\right)A^{d2}_{N,j}(t) + \xi_n A^{d2}_{D,j}(t) + \xi_d A^{d2}_{M,j}(t) + \phi\rho T^{d2}_{N,j}(t) \tag{59}$$

$$\frac{dS^{d2}_{N,j}(t)}{dt} = \sigma\psi E^{d2}_{N,j}(t) - \left(\mu + \frac{1}{\gamma}\right)S^{d2}_{N,j}(t) + \xi_n S^{d2}_{D,j}(t) + \xi_d S^{d2}_{M,j}(t) \tag{60}$$

$$\frac{dT^{d2}_{N,j}(t)}{dt} = \eta_j(t)A^{d2}_{N,j}(t) + \mu S^{d2}_{N,j}(t) - \left(\rho + \frac{1}{\gamma}\right)T^{d2}_{N,j}(t) + \xi_n T^{d2}_{D,j}(t) + \xi_d T^{d2}_{M,j}(t) + \omega_c\rho T^t_{N,j}(t) \tag{61}$$

b. For doxy-PEP (D):

$$\frac{dU_{D,j}(t)}{dt} = \eta_j(t)p_d U_{N,j}(t) + \rho(1-\omega_c)\left(T^0_{D,j}(t) + T^t_{D,j}(t)\right) + \rho(1-\phi)\left(T^c_{D,j}(t) + T^{d2}_{D,j}(t)\right)$$
$$+\nu\left(A^0_{D,j}(t) + A^c_{D,j}(t) + A^t_{D,j}(t) + A^{d2}_{D,j}(t)\right) - ((1-e_d){\lambda_j}^0(t) + (1-e_d)f_c{\lambda_j}^c(t) + f_t{\lambda_j}^t(t) + f_{d2}{\lambda_j}^{d2}(t)$$
$$+ \xi_n + \eta_j(t)p_v + \frac{1}{\gamma})U_{D,j}(t) + \xi_d U_{V,j}(t) \tag{62}$$

$$\frac{dE^0_{D,j}(t)}{dt} = (1-e_d){\lambda_j}^0(t)U_{D,j}(t) - \left(\sigma + \xi_n + \frac{1}{\gamma}\right)E^0_{D,j}(t) + \xi_d E^0_{V,j}(t) \tag{63}$$

$$\frac{dA^0_{D,j}(t)}{dt} = (1-w_t)\sigma(1-\psi)E^0_{D,j}(t) - \left(\nu + \eta_j(t) + \xi_n + \frac{1}{\gamma}\right)A^0_{D,j}(t) + \xi_d A^0_{V,j}(t) \tag{64}$$

$$\frac{dS^0_{D,j}(t)}{dt} = (1-w_t)\sigma\psi E^0_{D,j}(t) - \left(\mu + \xi_n + \frac{1}{\gamma}\right)S^0_{D,j}(t) + \xi_d S^0_{V,j}(t) \tag{65}$$

$$\frac{dT^0_{D,j}(t)}{dt} = \eta_j(t)A^0_{D,j}(t) + \mu S^0_{D,j}(t) - \left(\rho + \xi_n + \frac{1}{\gamma}\right)T^0_{D,j}(t) + \xi_d T^0_{V,j}(t) \tag{66}$$

$$\frac{dE^c_{D,j}(t)}{dt} = (1-e_d)f_c{\lambda_j}^c(t)U_{D,j}(t) - \left(\sigma + \xi_n + \frac{1}{\gamma}\right)E^c_{D,j}(t) + \xi_d E^c_{V,j}(t) \tag{67}$$

$$\frac{dA^c_{D,j}(t)}{dt} = (1-w_t)\sigma(1-\psi)E^c_{D,j}(t) - \left(\nu + \eta_j(t) + \xi_n + \frac{1}{\gamma}\right)A^c_{D,j}(t) + \xi_d A^c_{V,j}(t) + \phi\rho T^c_{D,j}(t) \tag{68}$$

$$\frac{dS^c_{D,j}(t)}{dt} = (1-w_t)\sigma\psi E^c_{D,j}(t) - \left(\mu + \xi_n + \frac{1}{\gamma}\right)S^c_{D,j}(t) + \xi_d S^c_{V,j}(t) \tag{69}$$

$$\frac{dT^c_{D,j}(t)}{dt} = \eta_j(t)A^c_{D,j}(t) + \mu S^c_{D,j}(t) - \left(\rho + \xi_n + \frac{1}{\gamma}\right)T^c_{D,j}(t) + \xi_d T^c_{V,j}(t) + \omega_c\rho T^0_{D,j}(t) \tag{70}$$

$$\frac{dE^t_{D,j}(t)}{dt} = f_t{\lambda_j}^t(t)U_{D,j}(t) - \left(\sigma + \xi_n + \frac{1}{\gamma}\right)E^t_{D,j}(t) + \xi_d E^t_{V,j}(t) \tag{71}$$

$$\frac{dA^t_{D,j}(t)}{dt} = w_t\sigma(1-\psi)E^0_{D,j}(t) + \sigma(1-\psi)E^t_{D,j}(t) - \left(\nu + \eta_j(t) + \xi_n + \frac{1}{\gamma}\right)A^t_{D,j}(t) + \xi_d A^t_{V,j}(t) \tag{72}$$

$$\frac{dS^t_{D,j}(t)}{dt} = w_t\sigma\psi E^0_{D,j}(t) + \sigma\psi E^t_{D,j}(t) - \left(\mu + \xi_n + \frac{1}{\gamma}\right)S^t_{D,j}(t) + \xi_d S^t_{V,j}(t) \tag{73}$$

$$\frac{dT^t_{D,j}(t)}{dt} = \eta_j(t)A^t_{D,j}(t) + \mu S^t_{D,j}(t) - \left(\rho + \xi_n + \frac{1}{\gamma}\right)T^t_{D,j}(t) + \xi_d T^t_{V,j}(t) \tag{74}$$

$$\frac{dE^{d2}_{D,j}(t)}{dt} = f_{d2}{\lambda_j}^{d2}(t)U_{D,j}(t) - \left(\sigma + \xi_n + \frac{1}{\gamma}\right)E^{d2}_{D,j}(t) + \xi_d E^{d2}_{V,j}(t) \tag{75}$$

$$\frac{dA^{d2}_{D,j}(t)}{dt} = w_t\sigma(1-\psi)E^c_{D,j}(t) + \sigma(1-\psi)E^{d2}_{D,j}(t) - \left(\nu + \eta_j(t) + \xi_n + \frac{1}{\gamma}\right)A^{d2}_{D,j}(t)$$
$$+ \xi_d A^{d2}_{V,j}(t) + \phi\rho T^{d2}_{D,j}(t) \tag{76}$$

$$\frac{dS^{d2}_{D,j}(t)}{dt} = w_t\sigma\psi E^c_{D,j}(t) + \sigma\psi E^{d2}_{D,j}(t) - \left(\mu + \xi_n + \frac{1}{\gamma}\right)S^{d2}_{D,j}(t) + \xi_d S^{d2}_{V,j}(t) \tag{77}$$

$$\frac{dT^{d2}_{D,j}(t)}{dt} = \eta_j(t)A^{d2}_{D,j}(t) + \mu S^{d2}_{D,j}(t) - \left(\rho + \xi_n + \frac{1}{\gamma}\right)T^t_{D,j}(t) + \xi_d T^t_{V,j}(t) + \omega_c\rho T^t_{D,j}(t) \tag{78}$$

c. For doxy-PEP + vaccination (V):

$$\frac{dU_{V,j}(t)}{dt} = \eta_j(t)p_v U_{D,j}(t) + \eta_j(t)p_d U_{M,j}(t) + \rho(1-\omega_c)\left(T^0_{V,j}(t) + T^t_{V,j}(t)\right)$$
$$+\rho(1-\phi)\left(T^c_{V,j}(t) + T^{d2}_{V,j}(t)\right) + \nu\left(A^0_{V,j}(t) + A^c_{V,j}(t) + A^t_{V,j}(t) + A^{d2}_{V,j}(t)\right)$$
$$-((1-e_{vd}){\lambda_j}^0(t) + (1-e_{vd})f_c{\lambda_j}^c(t) + (1-e_v)f_t{\lambda_j}^t(t) + (1-e_v)f_{d2}{\lambda_j}^{d2}(t) + \xi_n + \xi_d + \frac{1}{\gamma})U_{V,j}(t) \tag{79}$$

$$\frac{dE^0_{V,j}(t)}{dt} = (1-e_{vd}){\lambda_j}^0(t)U_{V,j}(t) - \left(\sigma + \xi_n + \xi_d + \frac{1}{\gamma}\right)E^0_{V,j}(t) \tag{80}$$

$$\frac{dA_{V,j}^0(t)}{dt} = (1-w_t)\sigma(1-\psi)E_{V,j}^0(t) - \left(\nu + \eta_j(t) + \xi_n + \xi_d + \frac{1}{\gamma}\right)A_{V,j}^0(t) \tag{81}$$

$$\frac{dS_{V,j}^0(t)}{dt} = (1-w_t)\sigma\psi E_{V,j}^0(t) - \left(\mu + \xi_n + \xi_d + \frac{1}{\gamma}\right)S_{V,j}^0(t) \tag{82}$$

$$\frac{dT_{V,j}^0(t)}{dt} = \eta_j(t)A_{V,j}^0(t) + \mu S_{V,j}^0(t) - \left(\rho + \xi_n + \xi_d + \frac{1}{\gamma}\right)T_{V,j}^0(t) \tag{83}$$

$$\frac{dE_{V,j}^c(t)}{dt} = (1-e_{vd})f_c\lambda_j{}^c(t)U_{V,j}(t) - \left(\sigma + \xi_n + \xi_d + \frac{1}{\gamma}\right)E_{V,j}^c(t) \tag{84}$$

$$\frac{dA_{V,j}^c(t)}{dt} = (1-w_t)\sigma(1-\psi)E_{V,j}^c(t) - \left(\nu + \eta_j(t) + \xi_n + \xi_d + \frac{1}{\gamma}\right)A_{V,j}^c(t) + \phi\rho T_{V,j}^c(t) \tag{85}$$

$$\frac{dS_{V,j}^c(t)}{dt} = (1-w_t)\sigma\psi E_{V,j}^c(t) - \left(\mu + \xi_n + \xi_d + \frac{1}{\gamma}\right)S_{V,j}^c(t) \tag{86}$$

$$\frac{dT_{V,j}^c(t)}{dt} = \eta_j(t)A_{V,j}^c(t) + \mu S_{V,j}^c(t) - \left(\rho + \xi_n + \xi_d + \frac{1}{\gamma}\right)T_{V,j}^c(t) + \omega_c\rho T_{V,j}^0(t) \tag{87}$$

$$\frac{dE_{V,j}^t(t)}{dt} = (1-e_v)f_t\lambda_j{}^t(t)U_{V,j}(t) - \left(\sigma + \xi_n + \xi_d + \frac{1}{\gamma}\right)E_{V,j}^t(t) \tag{88}$$

$$\frac{dA_{V,j}^t(t)}{dt} = w_t\sigma(1-\psi)E_{V,j}^0(t) + \sigma(1-\psi)E_{V,j}^t(t) - \left(\nu + \eta_j(t) + \xi_n + \xi_d + \frac{1}{\gamma}\right)A_{V,j}^t(t) \tag{89}$$

$$\frac{dS_{V,j}^t(t)}{dt} = w_t\sigma\psi E_{V,j}^0(t) + \sigma\psi E_{V,j}^t(t) - \left(\mu + \xi_n + \xi_d + \frac{1}{\gamma}\right)S_{V,j}^t(t) \tag{90}$$

$$\frac{dT_{V,j}^t(t)}{dt} = \eta_j(t)A_{V,j}^t(t) + \mu S_{V,j}^t(t) - \left(\rho + \xi_n + \xi_d + \frac{1}{\gamma}\right)T_{V,j}^t(t) \tag{91}$$

$$\frac{dE_{V,j}^{d2}(t)}{dt} = (1-e_v)f_{d2}\lambda_j{}^{d2}(t)U_{V,j}(t) - \left(\sigma + \xi_n + \xi_d + \frac{1}{\gamma}\right)E_{V,j}^{d2}(t) \tag{92}$$

$$\frac{dA_{V,j}^{d2}(t)}{dt} = w_t\sigma(1-\psi)E_{V,j}^c(t) + \sigma(1-\psi)E_{V,j}^{d2}(t) - \left(\nu + \eta_j(t) + \xi_n + \xi_d + \frac{1}{\gamma}\right)A_{V,j}^{d2}(t) + \phi\rho T_{V,j}^{d2}(t) \tag{93}$$

$$\frac{dS_{V,j}^{d2}(t)}{dt} = w_t\sigma\psi E_{V,j}^c(t) + \sigma\psi E_{V,j}^{d2}(t) - \left(\mu + \xi_n + \xi_d + \frac{1}{\gamma}\right)S_{V,j}^{d2}(t) \tag{94}$$

$$\frac{dT_{V,j}^{d2}(t)}{dt} = \eta_j(t)A_{V,j}^{d2}(t) + \mu S_{V,j}^{d2}(t) - \left(\rho + \xi_n + \xi_d + \frac{1}{\gamma}\right)T_{V,j}^t(t) + \omega_c\rho T_{V,j}^t(t) \tag{95}$$

d. For vaccination (M):

$$\begin{aligned}\frac{dU_{M,j}(t)}{dt} &= \eta_j(t)p_vU_{N,j}(t) + \xi_nU_{V,j}(t) + \rho(1-\omega_c)\left(T_{M,j}^0(t) + T_{M,j}^t(t)\right) \\ &+\rho(1-\phi)\left(T_{M,j}^c(t) + T_{M,j}^{d2}(t)\right) + \nu\left(A_{M,j}^0(t) + A_{M,j}^c(t) + A_{M,j}^t(t) + A_{M,j}^{d2}(t)\right) \\ &-\begin{pmatrix}(1-e_v)\lambda_j{}^0(t) + (1-e_v)f_c\lambda_j{}^c(t) + (1-e_v)f_t\lambda_j{}^t(t) \\ +(1-e_v)f_{d2}\lambda_j{}^{d2}(t) + \eta_j(t)p_d + \xi_d + \frac{1}{\gamma}\end{pmatrix}U_{M,j}(t)\end{aligned} \tag{96}$$

$$\frac{dE_{M,j}^0(t)}{dt} = (1-e_v)\lambda_j{}^0(t)U_{M,j}(t) - \left(\sigma + \frac{1}{\gamma}\right)E_{M,j}^0(t) + \xi_nE_{V,j}^0(t) - \xi_dE_{M,j}^0(t) \tag{97}$$

$$\frac{dA_{M,j}^0(t)}{dt} = \sigma(1-\psi)E_{M,j}^0(t) - \left(\nu + \eta_j(t) + \frac{1}{\gamma}\right)A_{M,j}^0(t) + \xi_nA_{V,j}^0(t) - \xi_dA_{M,j}^0(t) \tag{98}$$

$$\frac{dS_{M,j}^0(t)}{dt} = \sigma\psi E_{M,j}^0(t) - \left(\mu + \frac{1}{\gamma}\right)S_{M,j}^0(t) + \xi_nS_{V,j}^0(t) - \xi_dS_{M,j}^0(t) \tag{99}$$

$$\frac{dT_{M,j}^0(t)}{dt} = \eta_j(t)A_{M,j}^0(t) + \mu S_{M,j}^0(t) - \left(\rho + \frac{1}{\gamma}\right)T_{M,j}^0(t) + \xi_nT_{V,j}^0(t) - \xi_dT_{M,j}^0(t) \tag{100}$$

$$\frac{dE_{M,j}^c(t)}{dt} = (1-e_v)f_c\lambda_j{}^c(t)U_{M,j}(t) - \left(\sigma + \frac{1}{\gamma}\right)E_{M,j}^c(t) + \xi_nE_{V,j}^c(t) - \xi_dE_{M,j}^c(t) \tag{101}$$

$$\frac{dA_{M,j}^{c}(t)}{dt} = \sigma(1-\psi)E_{M,j}^{c}(t) - \left(\nu + \eta_j(t) + \frac{1}{\gamma}\right)A_{M,j}^{c}(t) + \xi_n A_{V,j}^{c}(t) - \xi_d A_{M,j}^{c}(t) + \phi\rho T_{M,j}^{c}(t) \quad (102)$$

$$\frac{dS_{M,j}^{c}(t)}{dt} = \sigma\psi E_{M,j}^{c}(t) - \left(\mu + \frac{1}{\gamma}\right)S_{M,j}^{c}(t) + \xi_n S_{V,j}^{c}(t) - \xi_d S_{M,j}^{c}(t) \quad (103)$$

$$\frac{dT_{M,j}^{c}(t)}{dt} = \eta_j(t)A_{M,j}^{c}(t) + \mu S_{M,j}^{c}(t) - \left(\rho + \frac{1}{\gamma}\right)T_{M,j}^{c}(t) + \xi_n T_{V,j}^{c}(t) - \xi_d T_{M,j}^{c}(t) + \omega_c\rho T_{M,j}^{0}(t) \quad (104)$$

$$\frac{dE_{M,j}^{t}(t)}{dt} = (1-e_v)f_t\lambda_j{}^{t}(t)U_{M,j}(t) - \left(\sigma + \frac{1}{\gamma}\right)E_{M,j}^{t}(t) + \xi_n E_{V,j}^{t}(t) - \xi_d E_{M,j}^{t}(t) \quad (105)$$

$$\frac{dA_{M,j}^{t}(t)}{dt} = \sigma(1-\psi)E_{M,j}^{t}(t) - \left(\nu + \eta_j(t) + \frac{1}{\gamma}\right)A_{M,j}^{t}(t) + \xi_n A_{V,j}^{t}(t) - \xi_d A_{M,j}^{t}(t) \quad (106)$$

$$\frac{dS_{M,j}^{t}(t)}{dt} = \sigma\psi E_{M,j}^{t}(t) - \left(\mu + \frac{1}{\gamma}\right)S_{M,j}^{t}(t) + \xi_n S_{V,j}^{t}(t) - \xi_d S_{M,j}^{t}(t) \quad (107)$$

$$\frac{dT_{M,j}^{t}(t)}{dt} = \eta_j(t)A_{M,j}^{t}(t) + \mu S_{M,j}^{t}(t) - \left(\rho + \frac{1}{\gamma}\right)T_{M,j}^{t}(t) + \xi_n T_{V,j}^{t}(t) - \xi_d T_{M,j}^{t}(t) \quad (108)$$

$$\frac{dE_{M,j}^{d2}(t)}{dt} = (1-e_v)f_{d2}\lambda_j{}^{d2}(t)U_{M,j}(t) - \left(\sigma + \frac{1}{\gamma}\right)E_{M,j}^{d2}(t) + \xi_n E_{V,j}^{d2}(t) - \xi_d E_{M,j}^{d2}(t) \quad (109)$$

$$\frac{dA_{M,j}^{d2}(t)}{dt} = \sigma(1-\psi)E_{M,j}^{d2}(t) - \left(\nu + \eta_j(t) + \frac{1}{\gamma}\right)A_{M,j}^{d2}(t) + \xi_n A_{V,j}^{d2}(t) - \xi_d A_{M,j}^{d2}(t) + \phi\rho T_{M,j}^{d2}(t) \quad (110)$$

$$\frac{dS_{M,j}^{d2}(t)}{dt} = \sigma\psi E_{M,j}^{d2}(t) - \left(\mu + \frac{1}{\gamma}\right)S_{M,j}^{d2}(t) + \xi_n S_{V,j}^{d2}(t) - \xi_d S_{M,j}^{d2}(t) \quad (111)$$

$$\frac{dT_{M,j}^{d2}(t)}{dt} = \eta_j(t)A_{M,j}^{d2}(t) + \mu S_{M,j}^{d2}(t) - \left(\rho + \frac{1}{\gamma}\right)T_{M,j}^{d2}(t) + \xi_n T_{V,j}^{d2}(t) - \xi_d T_{M,j}^{d2}(t) + \omega_c\rho T_{M,j}^{t}(t) \quad (112)$$

#### 3.3. Simulations

Posterior estimates of the transition parameters (model parameters were calibrated using Markov chain Monte Carlo (MCMC) to obtain the joint posterior distribution as detailed in Section 2; Table 3) were used to simulate disease dynamics - with and without intervention strategies - using the deSolve package (version 1.40). To quantify uncertainty, 1,000 parameter sets were sampled from this posterior and used in forward simulations. This procedure propagates uncertainty through the model by exploring the full joint posterior, thereby capturing variability in outcomes as well as parameter correlations. Sensitivity analyses were additionally performed for parameters not estimated through calibration, including uptake rates of doxy-PEP and vaccination. Specifically, simulations were conducted over time from 2027 to 2041 for MSM populations in England. Two alternative future scenarios were considered: (A) a stabilized behaviour scenario, in which the time-varying behavioural parameters inferred from historical data (i.e., force of infection $\lambda_j(t)$ and asymptomatic screening rate $\eta_j(t)$) remain constant beyond the final year where data was available; and (B) a continued trend scenario, where these behavioural trends continue to evolve through to 2041 based on past incidence.

Moreover, rather than tracking clinically manifest cases, this modelling framework quantifies the incidence of newly acquired infections, defined explicitly as the biological event of successful pathogen invasion and replication within a host. This metric comprehensively captures all individuals entering the Exposed $E$ compartment, encompassing the entire cohort of hosts harbouring *N. gonorrhoeae* irrespective of symptomatic presentation or subclinical status (i.e., both symptomatic and asymptomatic infections). For each calendar year $t$, the total number of gonorrhoea infection $Y^k(t)$ across all strata $i \in \{N, X, D, M\}$ for strains $k \in \{0, c, t, d2\}$, are as follows:

$$Y^0(t) = \sum_{j\in\{L,H\}} \left( \int_t^{t+1} \lambda_j^0 \big(U_{N,j} + (1-e_d)U_{D,j} + (1-e_{vd})U_{V,j} + (1-e_v)U_{M,j}\big)(\tau)d\tau \right) \quad (113)$$

$$Y^c(t) = \sum_{j\in\{L,H\}} \left( \int_t^{t+1} \lambda_j^c \big(f_c U_{N,j} + (1-e_d) f_c U_{D,j} + (1-e_{vd}) f_c U_{V,j} + (1-e_v) f_c U_{M,j}\big)(\tau) d\tau \right) \tag{114}$$

$$Y^t(t) = \sum_{j\in\{L,H\}} \left( \int_t^{t+1} \lambda_j^t \big(f_t U_{N,j} + f_t U_{D,j} + (1-e_v) f_t U_{V,j} + (1-e_v) f_t U_{M,j}\big)(\tau) d\tau \right) \tag{115}$$

$$Y^{d2}(t) = \sum_{j\in\{L,H\}} \left( \int_t^{t+1} \lambda_j^{d2} \big(f_{d2} U_{N,j} + f_{d2} U_{D,j} + (1-e_v) f_{d2} U_{V,j} + (1-e_v) f_{d2} U_{M,j}\big)(\tau) d\tau \right) \tag{116}$$

with $\tau$ denotes the time variable of integration.

**Section 4: Supplementary results**

**4.1. Performance metrics**

To evaluate the emergence of different gonorrhoea strains $k \in \{0, c, t, d2\}$, we compare the total number of gonorrhoea infections $Y^k(t)$ to the corresponding baseline without intervention, $\hat{Y}^k(t)$, to calculate the total number of averted gonorrhoea infections over $M$ years (starting from year $t_0$):

$$\sum_{t=1}^{M} [\hat{Y}^k(t_0+t) - Y^k(t_0+t)] \tag{117}$$

as well as in terms of percentage decrease:

$$\frac{\sum_{t=1}^{M} [\hat{Y}^k(t_0+t) - Y^k(t_0+t)]}{\sum_{t=1}^{M} [\hat{Y}^k(t_0+t)]} \times 100\% \tag{118}$$

Furthermore, to evaluate whether a given intervention configuration establishes long-term strain-specific stabilization or prompts a complete ecological displacement toward multidrug-resistant lineages, we quantify relative strain composition at the terminal horizon of the simulation period (year $M$) for each strain $k \in \{0, c, t, d2\}$, respectively:

$$\frac{Y^k(M)}{\sum_{i\in\{0,c,t,d2\}} Y^i(M)} \tag{119}$$

In addition, to evaluate the programmatic efficiency of each intervention, we calculate the total number of doxy-PEP enrolments and/or the total number of vaccine doses administered over $M$ years (starting from year $t_0$):

$$\sum_{t=1}^{M} [p_d(Y_U^N(t_0+t) + Y_U^M(t_0+t)) + p_v(Y_U^N(t_0+t) + Y_U^D(t_0+t))] \tag{120}$$

where

$$Y_U^i(t) = \sum_{j\in\{L,H\}} \left( \int_t^{t+1} \eta_j(\tau) U_j^i(\tau) d\tau \right) \tag{121}$$

Lastly, for each gonorrhoea strains $k \in \{0, c, t, d2\}$, we calculate the number of gonorrhoea infections averted per intervention programme enrolment over a period of $M$ years, beginning from year $t_0$:

$$\frac{\sum_{t=1}^{M}[\hat{Y}^k(t_0+t)-Y^k(t_0+t)]}{\sum_{t=1}^{M}[p_d(Y_U^N(t_0+t)+Y_U^M(t_0+t))+p_v(Y_U^N(t_0+t)+Y_U^D(t_0+t))]} \tag{122}$$

Here, an "enrolment" is defined operationally as an eligible individual accepting the offer of doxy-PEP and/or vaccination at a sexual health clinic, rather than tracking the precise quantity of doxycycline and/or vaccine doses subsequently consumed.

**4.2. Results of increasing transmissibility of gonorrhoea**

To further examine the robustness of these interventions, we performed a sensitivity analysis under the assumption of continuously increasing transmissibility of gonorrhoea within the MSM population. To parameterize the forward-looking projection window, the model was initialized in calendar year 2027. Reflecting low-level cryptic circulation and documented global importation, the initial population-level strain-specific resistance proportions within the infected reservoir were established at $1 \times 10^{-4}$ for the Cef-R strain and $8 \times 10^{-5}$ for Dual-R strain [19]. Uncertainty across all metrics was propagated from the joint posterior distribution and reported as median estimates with 95% credible intervals (CrI).

Assuming a fixed 66% intervention uptake rate within the target population, and incorporating an empirically calibrated, near-zero ceftriaxone therapeutic treatment failure rate $\phi$ derived from historical national surveillance data, this scenario significantly alters the long-term effectiveness of both pharmacological and immunological interventions, as the rising baseline transmission intensity competes with the protective effects of the rollout (see Fig. 5). Under this scenario, the total burden of gonorrhoea infections in the no-intervention scenario exhibited a sustained upward trajectory. While doxy-PEP standalone intervention also showed an increase in total infections, it maintains a marginally lower incidence than the baseline. Conversely, both vaccination and the combined strategy initially drive a reduction in total cases; however, this trend eventually reversed into a "rebound" phase as the increase in behaviour and the waning of vaccine-induced immunity converge to drive a late-stage surge in transmission. The dynamics of individual strains under increased behavioural pressure reveal distinct evolutionary pathways. For baseline susceptible strains, the no-intervention baseline reflects the broader population trend of increasing incidence. Vaccination initially suppresses these infections, but a subsequent increase is observed as immunity wanes. Notably, both doxy-PEP and the combined strategy remain highly effective at driving baseline susceptible strains toward elimination, with the synergistic effect of the combination strategy accelerating this clearance despite the heightened transmission background. Similarly, for Cef-R strains, all strategies facilitate a decline, but doxy-PEP and the combined strategy are uniquely capable of achieving complete elimination, outperforming both the no-intervention and vaccination-only scenarios. Tet-R strains exhibit the most significant variation under increased behavioural assumptions. While the no-intervention baseline for tetracycline-resistant gonorrhoea stabilizes, suggesting a balance between increased transmission and fitness costs, doxy-PEP standalone intervention drives a continuous and aggressive expansion of this resistant population. The combined strategy initially suppresses Tet-R infections to levels below the baseline; however, following the waning of vaccine protection, the underlying selective pressure of doxy-PEP causes Tet-R infections to surge, eventually exceeding the absolute number seen in the no-intervention scenario. Dual-resistant strains, by contrast, continue to decline across all scenarios, though the rate of reduction is notably slowest under doxy-PEP standalone intervention and most rapid following vaccination.

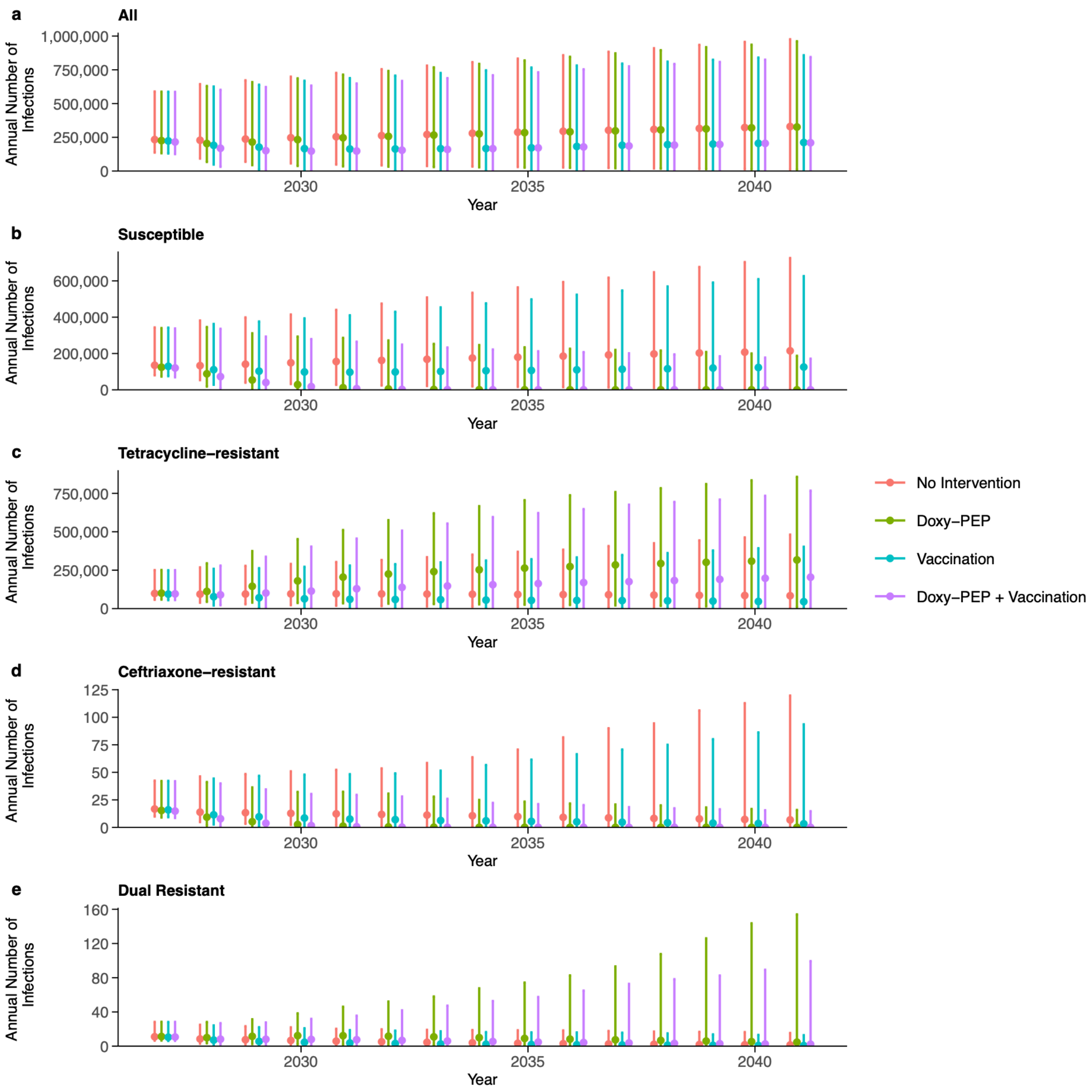


**Fig. 5: Simulated long-term impact of doxy-PEP and moderate-efficacy vaccination on *N. gonorrhoeae* transmission volumes and resistance dynamics under baseline treatment efficacy**. For each point plot, dots denote median estimates, and the accompanying vertical error bars represent the corresponding 95% CrI. Projections span a 15-year horizon (2027-2041) under the continued behavioural baseline (Scenario B), assuming a fixed 66% intervention uptake rate within the target population. Model simulations incorporate an empirically calibrated, near-zero ceftriaxone therapeutic treatment failure rate $\phi$ derived from historical national surveillance data. Forward projections are initialized in 2027 with population-level strain-specific resistance proportions established at $1 \times 10^{-4}$ for Cef-R and $8 \times 10^{-5}$ for Dual-R strains. Subpanels depict trajectories for total incident infections alongside disaggregated strain-specific volumes for baseline susceptible, Tet-R, Cef-R, and Dual-R strains.

Next, to evaluate model robustness against escalating AMR selection, under an assumed 66% intervention uptake within the target population, a stress-test sensitivity analysis was conducted by artificially elevating the ceftriaxone therapeutic treatment failure parameter $\phi$ to 20%, simulating a profound compromise in frontline cephalosporin efficacy (Fig. 6). Under this high-failure scenario, the transmission trajectories for aggregate incident infections, baseline susceptible strains, and Tet-R lineages exhibit epidemiological dynamics that are qualitatively identical to those observed in the near-zero baseline treatment failure scenario. The defining departure from the primary analysis occurs within the Cef-R and Dual-R strain populations (Fig. 6d and Fig. 6e).

At a 20% treatment failure rate, the unexposed no-intervention scenario crosses a critical epidemiological threshold, shifting from a baseline downward trajectory toward a sustained, linear increase with significant outbreak potential, reaching 660 (95% CrI: 12 – 323900) annual incident infections by year 15. Crucially, while the doxy-PEP standalone strategy, vaccination standalone strategy, and the combined dual-intervention strategy all remain effective at suppressing the standalone Cef-R strain, averting a cumulative total of 740 (95% CrI: 20 – 1237300) infections, 2500 (95% CrI: 130 – 1451500) infections, and 2530 (95% CrI: 130 – 1451500) infections, respectively. Under the doxy-PEP standalone intervention, the Dual-R population has the probability to undergo aggressive epidemic expansion to 400 (95% CrI: 6 – 365600) infections by year 15, presenting a substantial outbreak trajectory that exceeds even the unexposed baseline of 110 (95% CrI: 3 – 71400) infections by the 2041 horizon, yielding a negative cumulative total of -1160 (95% CrI: -1094700 – -45) averted infections across the 15-year horizon. In stark contrast, vaccination alone and the combined strategy can maintain suppression of both Cef-R and Dual-R lineages. For the Cef-R strain, the vaccination standalone strategy averts a cumulative total of 740 (95% CrI: 20 - 1237300) infections, while the combined strategy averts 2530 (95% CrI: 130 – 1451500) total infections. For the Dual-R strain, the vaccination standalone strategy averts a cumulative total of 180 (95% CrI: 8 – 261300) infections, whereas the combined strategy yields a slightly negative prophylactic balance of -140 (95% CrI: -123900 – 148700) infections. By year 15, Cef-R annual incidence increases to 150 (95% CrI: 0 – 80000) infections under vaccination standalone strategy compared with 0 (95% CrI: 0 – 90) infections under the combined strategy. Similarly, Dual-R annual incidence is restricted to 30 (95% CrI: 0 – 15200) infections under vaccination compared with 80 (95% CrI: 0 – 115500) infection under the dual-intervention approach.

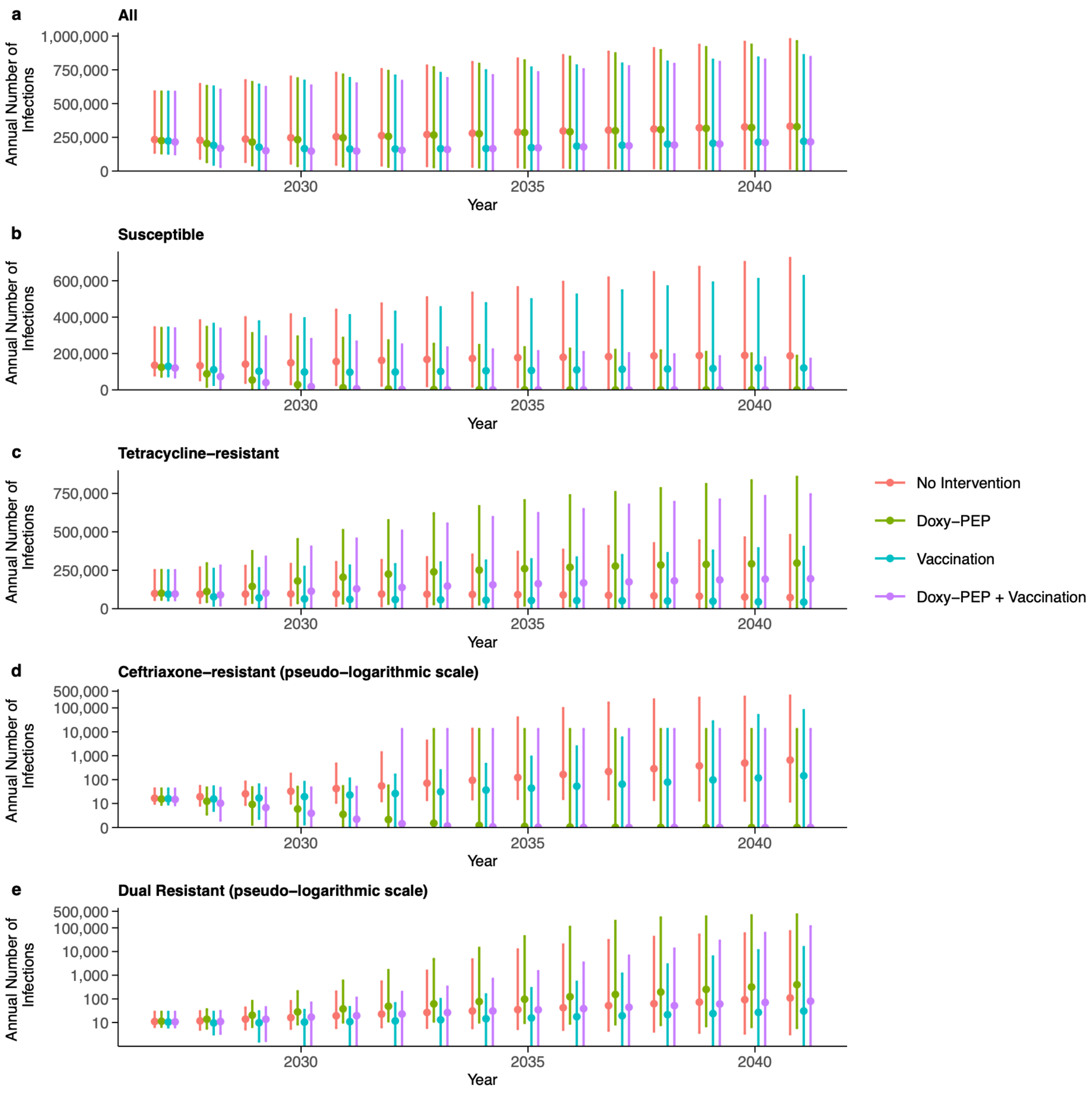


**Fig. 6: Simulated long-term impact of doxy-PEP and moderate-efficacy vaccination on *N. gonorrhoeae* transmission volumes and resistance dynamics under accelerated frontline treatment failure.** For each point plot, dots denote median estimates, and the accompanying vertical error bars represent the corresponding 95% CrI. Projections span a 15-year horizon (2027-2041) under the continued behavioural baseline (Scenario B), assuming a fixed 66% intervention uptake rate within the target population. Model simulations incorporate an artificially elevated ceftriaxone therapeutic treatment failure rate ($\phi = 20\%$) to evaluate system robustness against frontline cephalosporin compromise. Forward projections are initialized in 2027 with population-level strain-specific resistance proportions established at $1 \times 10^{-4}$ for Cef-R and $8 \times 10^{-5}$ for Dual-R strains. Subpanels depict trajectories for total incident infections alongside disaggregated strain-specific volumes for baseline susceptible, Tet-R, Cef-R, and Dual-R strains. To improve visualization across several orders of magnitude, the y-axes in panels d (Cef-R) and e (Dual-R) are displayed using a pseudo-logarithmic ($log_{10}$-equivalent) scale, whereas panels a-c use a linear scale.
